# Scaling Up the Discovery of Hesitancy Profiles by Identifying the Framing of Beliefs towards Vaccine Confidence in Twitter Discourse

**DOI:** 10.1101/2021.10.01.21264439

**Authors:** Maxwell A. Weinzierl, Suellen Hopfer, Sanda M. Harabagiu

**Author notes:** **Contributions** All authors contributed to the study conception and design. Material preparation and data collection was performed by Maxwell Weinzierl, and data analysis was performed by all authors. Multiple drafts of the manuscript were written by Maxwell Weinzierl, Sanda Harabagiu, and Suellen Hopfer, and all authors commented on previous versions of the manuscript. All authors read and approved the final manuscript. **Declarations**. **Ethics approval** All procedures performed in this study were in accordance with the ethical standards of the institutional and/or national research committee and with the 1964 Helsinki Declaration and its later amendments or comparable ethical standards. The study was approved by The University of Texas at Dallas Institutional Review Board (IRB-21-515 stipulated that our research met the criteria for exemption #8(iii) of the Chapter 45 of Federal Regulations Part 46.101.(b)). **Consent to participate** Informed consent was obtained from all individual participants included in the study. **Consent for publication** The authors affirm that research participants provided informed consent for publication of aggregated results.

## Abstract

Our study focused on the inference of the framing of confidence in the HPV vaccine throughout a collection of 422,078 tweets as well as the framing of confidence in the COVID-19 vaccines through a collection of 5,865,046 tweets. The vaccine confidence framings were inferred by using a novel Question/Answering framework enabling the derivation of a misinformation taxonomy as well as trust taxonomies for these two vaccines. These taxonomies, along with the analysis of vaccine literacy, the implied moral foundations and the tension between vaccine mandates and civil rights allowed us to discover several profiles of hesitancy for each vaccine across 138,779 Twitter users referring to confidence in HPV vaccine and 665,798 users referring to confidence in COVID-19 vaccines. These hesitancy profiles inform public health messaging approaches to effectively reach Twitter users with promise to shift or bolster vaccine attitudes.

## INTRODUCTION

Social media microblogging platforms, specifically Twitter, have become highly influential and relevant to shaping attitudes towards vaccination. With 206 million daily active users as of 2021, Twitter has substantial reach and daily exposure being the most popular social network for news consumption (Auxier & Anderson, 2021; Statista, 2021). Moreover, Twitter allows people to express their beliefs about vaccine confidence or hesitancy, their trust or mistrust in vaccines as well as their stance on civil rights and vaccination mandates. However, as reported in Karafillakis (2021), social media monitoring of vaccination discourse reported in literature typically involves the recognition of positive or negative sentiments (Du et al., 2017; Luo et al., 2019; Massey et al., 2016) towards several vaccination topics (Surian et al., 2016) or entities extracted from tweets (Luo et al., 2019), where vaccine hesitancy is associated with neutral sentiment.

In this study we take a fundamentally different view on the discovery of vaccine hesitancy in Twitter discourse, by relying on a Question/Answering (Q/A) framework that enables the inference of vaccine confidence framings. Our method is informed by the 2011 interdisciplinary World Health Organization’s Strategic Advisory Group of Experts (SAGE) Working Group on Vaccine Hesitancy definition of vaccine hesitancy as the “delay in acceptance or refusal of vaccines despite its availability” (MacDonald & group, 2015). The working group recognized at least three universal factors (3C Model) contributing to vaccine hesitancy, subsequently developing the 3C vaccine hesitancy model consisting of (1) vaccine confidence, (2) vaccine complacency, and (3) vaccine constraints (practical vaccine barriers). The 3C and subsequent vaccine hesitancy models (Betsch et al., 2018) have shown vaccine confidence to play a significant role and explain the most substantial proportion of variance underpinning vaccine doubt that in turn contributes to individuals not vaccinating. We therefore set out to examine the degree to which vaccine confidence was framed on social media and how it informs profiles of vaccine hesitancy for both HPV and COVID-19 - the two most controversial yet effective and underutilized vaccines for which there remains substantial reluctance among the public. Our first research question therefore asks:

### RQ1: How is confidence in the HPV and the COVID-19 vaccines framed in the Twitter discourse?

Answering this question was possible by casting the search for framings of vaccine confidence as a Twitter Question/Answering (Q/A) problem, using the questions introduced in (Rossen et al., 2019), as the Vaccine Confidence Repository (VCR). The VCR is informed by the antivaccine content analysis of Kata (Kata, 2010, 2012). In the study reported in Rossen (2019), the questions comprising the VCR were used as survey links available from Facebook pages and parenting forums. Nearly 300 Australian visitors answered the questions, enabling the discovery of hesitancy profiles. We were inspired by this work, and instead of soliciting answers from Twitter users, we decided to (a) automatically find tweets that answer the same questions; and (b) infer the confidence framings referred by the answers. Moreover, we were interested to extend our method by covering not only confidence in the HPV vaccine, but also in the newer COVID-19 vaccine. Not only were we able to discern the confidence in vaccines framings on Twitter, but we also analyzed these confidence framings and observed that they often relied on misinformation. We considered misinformation as any misconception, references to conspiracy theories, or any flawed reasoning. More importantly, it is interesting to uncover the specific misinformation that is related to vaccine confidence. This allowed us to address the second research question of this study:

### RQ2: What specific misinformation about the HPV and COVID-19 vaccines is propagated on Twitter?

Answering RQ2 entails discovering the specific misinformation that was unveiled by answering the questions from VCR, but also and importantly, the derivation of a taxonomy of misinformation that is used to frame confidence in the HPV or the COVID-19 vaccines. Misinformation has exploded on social media platforms such as Twitter (Cacciatore, 2021; Hou et al., 2021; Wawrzuta et al., 2021), but it is less known which misinformation themes are propagated and what concerns they address. Building a taxonomy of misinformation to uncover HPV and COVID-19 vaccine related misinformation is needed, against which inoculation interventions can be prepared. A growing literature suggests that vaccine acceptance depends to a large extent on public trust and related confidence in the safety and efficacy of vaccines (Larson et al., 2018; Latkin et al., 2021; Siegrist, 2021). To further understand the way in which trust in vaccines impacts vaccine confidence, we considered a third question in our study:

### RQ3: What trust issues are associated with the HPV and COVID-19 vaccines in Twitter conversations?

The multidimensional concept of trust involves not only trust in the vaccine, but also trust in the healthcare practitioners who administer vaccines, the healthcare systems, public health authorities and governments who advocate for vaccination. Trust is increasingly important especially in the context of high uncertainty for vaccine decision-making such as with the recent coronavirus pandemic, rapidly changing emerging science on what is known about Coronavirus as example, changing vaccine recommendations, growing science illiteracy, and the growing number of vaccines being recommended. Under these conditions of uncertainty, the public depends increasingly on the expertise, judgements, competency, and transparency in sharing what is known about vaccines. The case of trust and vaccination carries with it a history of vaccine development and missteps, but social movements and reactions render trust in vaccines highly variable and locally specific (Larson et al., 2016). In the context of our study, we explored trust erosion, or increase, in vaccination. The answer to RQ2 led to the development of two trust taxonomies for each vaccine, namely a taxonomy for trust building and a taxonomy for eroding trust. These taxonomies revealed a constellation of concerns that addressed several trust themes impacting confidence in vaccines. Interestingly, we found that many of the trust concerns that we discovered aligned with a multitude of definitions of trust, ranging from the individual level, e.g. trust involving the overall reluctance to obtain vaccination due to fear of side effects (Latkin et al., 2021), to societal and system levels of trust in science and public health authorities (Siegrist, 2021; Sutton et al., 2020). Explaining the differences in vaccine trust or in the tension between civil rights and vaccine mandates is made possible by considering the moral aspects of the framings. Consequently, we also addressed the research question:

### RQ4: What moral dimensions characterize the confidence in the HPV and COVID-19 vaccines on Twitter?

Because previous work in social psychology considered the Moral Foundations Theory (MFT) (Haidt & Graham, 2007; Haidt & Joseph, 2004) as a theoretical framework for analyzing moral framing, we used the same five key values of human morality, emerging from evolutionary, social, and cultural origins to encode all the confidence framings. These moral encoding proved to be very informative in the discovery of the hesitancy profiles based on vaccine confidence, as revealed by Twitter discourse. Ultimately, in this study we were most interested to answer the research question:

### RQ5: What hesitancy profiles can be discerned from Twitter for the HPV and COVID-19 vaccines?

Answering this question entails discovering how hundreds of thousands of Twitter users frame their confidence in vaccines and what stance they have towards these framings. This was possible because we had access to tweets discussing the HPV vaccine authored by 192,487 users and tweets discussing the COVID-19 vaccine authored by 2,268,358 users. However, through the Q/A method presented in this paper, we found that only 138,779 Twitter users addressed confidence in the HPV vaccine and only 665,798 users addressed confidence in the COVID-19 vaccines. Our hypothesis is that Twitter authors that frame their vaccine confidence in similar ways, with respect to their adoption or rejection of misinformation, their erosion or building trust in the vaccines, the vaccine literacy they have or lack, their stance on the respect of civil rights as well as their focus on certain moral foundations belong to the same hesitancy profile. To our knowledge, this is the first study that aims at the automatic discovery of hesitancy profiles at large scale, especially by using confidence framings as answers inferred from a set of questions. Our search is expanded to find all users that share each confidence framing, as well as their acceptance or rejection of the framing. We believe that this method generates profiles that provide a richer interpretation than those reported in Rossen (2019), which were based on answers provided by human participants that indicated their agreement with each VCR question. We instead cast the attitudes toward each question as a set of five attitude-generating questions, automatically created from the same VCR questions used in Rossen (2019), extended with the same number of questions covering hesitancy for the COVID-19 vaccine. Of note is that the answers were not provided by Twitter users that considered the questions, but by framings of vaccine confidence, inferred from tweets deemed relevant to each question. The fact that framings were further characterized by linking them to the misinformation or trust taxonomies while also considering the moral foundations they allowed us to interpret the hesitancy profiles against this rich characterization of vaccine confidence, instead of only relying on the quantified attitudes of answers to the VCR questions. We were pleasantly surprised by the hesitancy profiles revealed by the method presented in this paper, and the insightful interpretations that could be derived. We believe that these profiles identify where interventions can be delivered on the Twitter platforms, and most importantly, what vaccine hesitancy issues the interventions need to consider. Finally, as the method was successfully applied for two different vaccines, it highlights its portability for considering confidence across vaccines and deriving vaccine-specific hesitancy profiles.

## METHODS

### Overview of Methodology

The methodology that we employed for uncovering vaccine hesitancy profiles from the Twitter discourse addressing vaccine confidence is illustrated in Figure 1. The methodology uses a Question/Answering (Q/A) framework operating on either COVID-19 and the HPV vaccine discourse. First, we considered (a) a general question asking about confidence in the HPV or COVID-19 vaccine, i.e. Q1: “*How confident are you in the safety of the HPV/COVID-19 vaccine*?” and (b) a set of 18 questions from the Vaccine Confidence Repository (VCR), introduced by Rossen et al. (2019), which concerns five major belief themes resulting from the analysis of the content expressed in anti-vaccination sites (Kata, 2010, 2012). The question belief themes are: (T1) vaccines are unsafe and unnatural; (T2) vaccines are ineffective; (T3) there is redundant vaccination; (T4) parents should be free to choose whether or not to vaccinate their children and (T5) vaccination is a conspiracy. For each belief theme, three or four questions were formulated. As shown in Figure 1, for each question we automatically generated five attitude-invoking questions. Each of the resulting questions was processed by a relevance model against the index of tweets.

**Figure 1:**
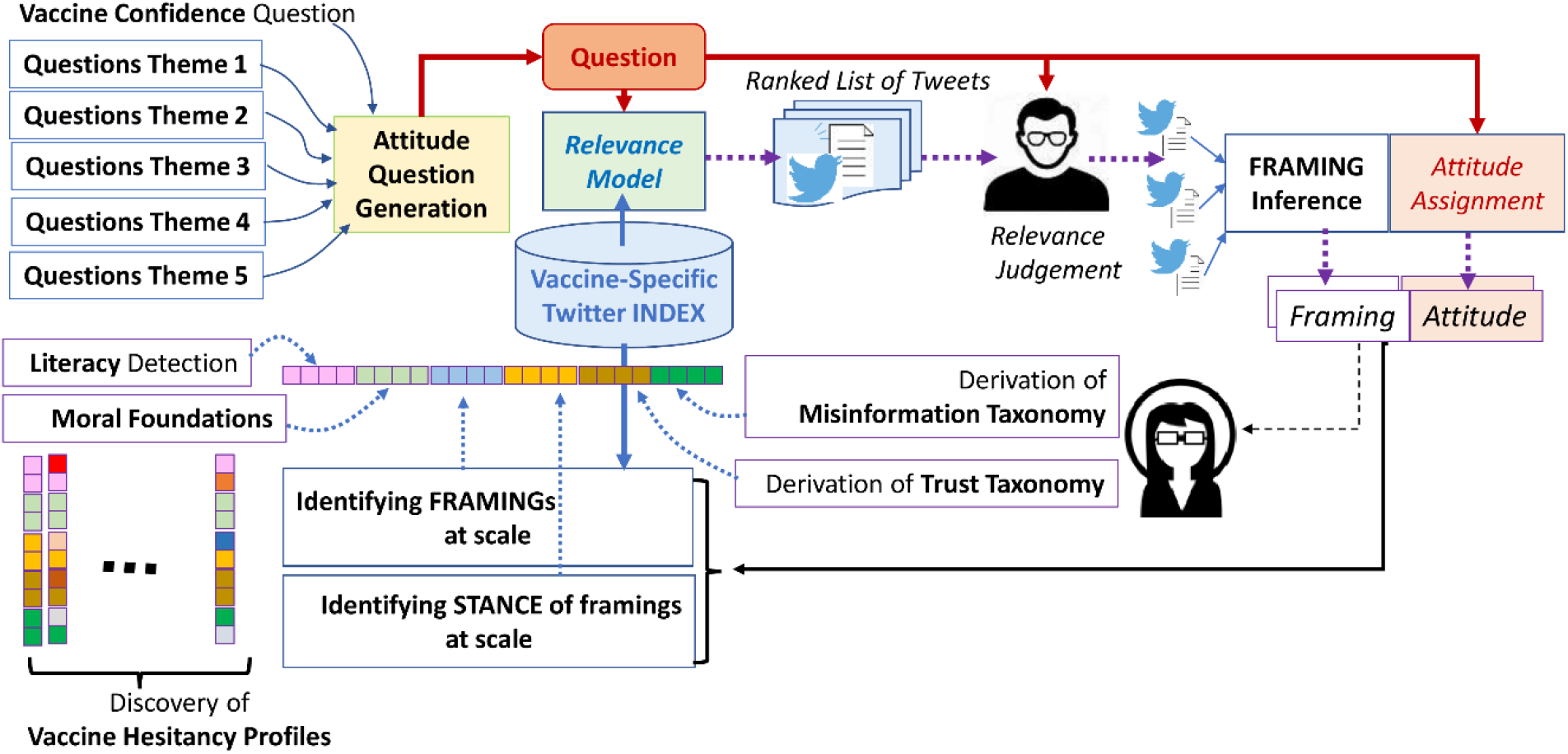
Uncovering the framing of attitudes towards vaccine confidence using a Question/Answering framework which leads to the derivation of vaccine hesitancy profiles, informed by Misinformation and Trust Taxonomies, Moral Foundations and vaccine literacy.

Table 1 lists one question from T2 (theme 2, vaccines are ineffective) pertaining to the quest for vaccine confidence in the COVID-19 vaccine along with the attitude-invoking questions generated from it, as well as a question from T2 used for inquiring about confidence in the HPV vaccine. Questions Q.C1-5 and Q.H1-5 were generated by using regular expressions operating on each of the initial 19 questions inquiring about confidence in each of the vaccines. In this way we generated 95 (19 × 5) attitude-evoking questions for each vaccine. Each of these 95 questions invoking attitudes were answered by retrieving a ranked list of relevant tweets, using a relevance model, implementing the BM25 vector ranking model (Beaulieu et al., 1997).

**Table 1:**
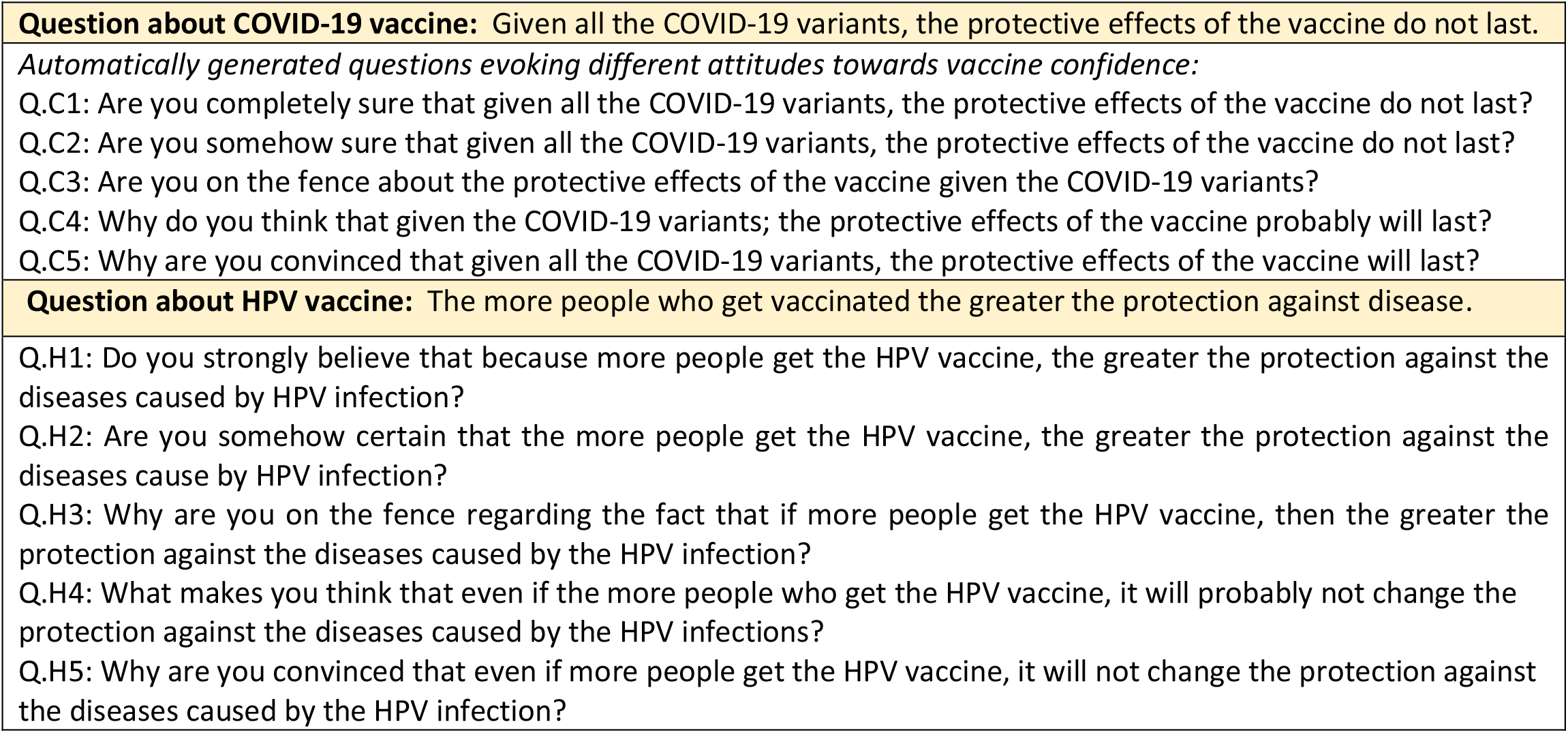
Examples of automatically generated attitude-evoking questions.

The collection that was searched for relevant tweets was obtained by using the Twitter streaming API for each vaccine. For the HPV vaccine, we used the Twitter historical API with the following query “(human papillomavirus vaccination) OR (human papillomavirus vaccine) OR gardasil OR cervarix OR (hpv vaccine) OR (hpv vaccination) OR (cervical vaccine) OR (cervical vaccination) lang:en”, 1,833,380 total tweets, with 969,372 retweets and 864,008 original tweets from 625,354 total authors. These tweets were authored in the time frame initiating on January 1^st^, 2008, and end ending on May 1^st^, 2021 (∼13 years). A large fraction of these tweets, which were duplicates likely due to spam bots, required filtering. Locality Sensitive Hashing (LSH) (Das et al., 2007) is a well-known method used to remove near-duplicate documents in large collections. We performed LSH, with term trigrams, 100 permutations, and a Jaccard threshold of 50%, on our original tweets collection to produce the collection 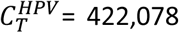 unique original tweets. The tweets from 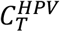 were authored by 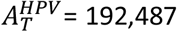 users. Using the same methodology, we used the query “(covid OR coronavirus) vaccine lang:en” for retrieving for the COVID-19 vaccines a collection of 19,021,575 total tweets, with 9,888,104 retweets and 9,133,471 original tweets from 4,382,289 total users obtained from the Twitter streaming API, which resulted after near-duplication removal into the collection 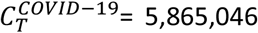 unique original tweets, authored by 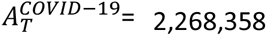 users in the time span January 17^th^, 2021 to July 21^st^, 2021 (∼6 months).

We used Lucene (lucene.apache.org) to index in *I*^*HPV*^ the tweets from 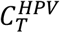 and in *I*^*COVID*-19^ the tweets from 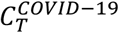. For each of the attitude-evoking questions pertaining to the confidence in the HPV vaccine, a set of ranked tweets from 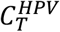 are retrieved. The ranking is produced by the scoring function from the BM25 relevance model, operating on the indexes *I*^*HPV*^ or *I*^*COVID*-19^. Similarly, for each of the attitude-evoking questions pertaining to the confidence in the COVID-19 vaccine, a set of ranked tweets from 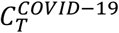 was retrieved. It is important to note that from the ranked list of tweets retrieved by the relevance mode, we considered (1) only the top 300 ranked tweets, and (2) we merged all the top ranked tweets retrieved for all attitude-evoking questions corresponding to any of the original 19 questions, aiming to judge their relevance to the question. The judgements were performed by two experts in question answering. A total of 1523 tweets were judged relevant to the questions by two researchers from the Human Language Technology Research Institute at [Blinded University]. Cohen’s Kappa score was 0.81, which indicates strong agreement between annotators (0.8-0.9) (Zapf et al., 2016).

As shown in Figure 1, in addition to assessing the relevance of tweets against questions, the human judges inferred framings used by Twitter users in the relevant tweets, to express their confidence in vaccines, as answers to the questions. The resulting sets of framings *framings*^*HPV*^ and *framings*^*COVID*-19^ inform the answer to RQ1. Additionally, the framings were also categorized as expressing (1) misinformation; (2) evoking issues of trust in vaccines; (3) pertaining to civil rights or (4) expressing morality issues. All framings expressing misinformation were used to derive a misinformation taxonomy, assigning ontological commitments for the misinformation *themes* and *concerns*. This allowed us to answer RQ2. Similarly, trust taxonomies were also derived by an ontology expert. This allowed us to answer RQ3. Furthermore, two experts assigned Moral Foundations, defined by the Moral Foundations Theory (MFT) (Haidt & Graham, 2007; Haidt & Joseph, 2004) to each framing from *framings*^*HPV*^ and *framings*^*COVID*-19^, informing our answer to RQ4. Furthermore, the framings from *framings*^*HPV*^ and *framings*^*COVID*-19^ that showcased vaccine literacy or lack of it were also annotated. Moreover, we wondered if there were more tweets in the 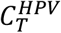 and in *C*^*COVID*-19^ collections that referred to the framings from *framings*^*HPV*^ and *framings*^*COVID*-19^. To find out, we automatically recognized these framings at scale, in our entire collections of tweets discussing COVID-19 or HPV vaccines. For this purpose, we relied on an automatic method, which was previously used to identify misinformation about the COVID-19 vaccine (Weinzierl & Harabagiu, 2021).

In addition, for each tweet from the 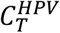 or 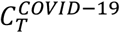 collections of tweets that referred to any of the framings from *framings*^*HPV*^ or *framings*^*COVID*-19^, we also automatically identified the stance of the tweet towards the framing, i.e. accepting it or refusing it, using the method reported in (Weinzierl et al., 2021). This allowed us to generate a representation of the users involved in the discourse about COVID-19 vaccines and the users participating in the discourse about the HPV vaccine by learning a vectorial user representation of (1) the framings the user evoked, (2) the stance the user had towards each framing; (3) the ontological commitments to misinformation and trust, provided by the taxonomies of misinformation and trust that we have derived; (4) the vaccine literacy of each user; (5) his/her beliefs about civil rights and vaccination, and (6) the moral foundations the user implied. The resulting vectorial user representations informed the discovery of vaccine hesitancy profiles in our Twitter datasets. Moreover, we were able to find the characteristics of each profile of vaccine hesitancy – by interpreting their predominant stance towards trust in vaccines, misinformation, civil rights, or moral foundations.

### Framing Vaccine Hesitancy Based on Beliefs in Vaccine Confidence

Tweets that: (1) are relevant to the same question and (2) share the same attitude in response to the question can be further segregated by similarity of their content and the way the authors’ viewpoint is framed. Table 2 shows how in this example, from all the six tweets that were judged to be against the predication of the question illustrated in the Table, Framing A was inferred. Similarly, the tweets judged as doubting or accepting the question predication are shown in Table 2, along with their framings. A total of 113 framings were inferred in *framings*^*COVID*-19^ and 64 framings in *framings*^*HPV*^.

**Table 2:** Example of question used to inquire about the confidence in COVID-19 vaccines, answered by relevant tweets. The question belongs to the theme of unsafe and unnatural vaccinations. The relevant tweets are categorized according to the attitudes expressed towards the question predication. Examples of inferred framings for each attitude category are shown.

|  |
| --- |
| **Q(COVID-19):** *Vaccines have not been adequately tested for safety.*<br><u>**Tweets AGAINST the question predication:**</u><br><u>**Tweet 1:**</u> Scientists have been working on Coronavirus vaccines for decades; once COVID-19 was isolated they could focus attention on that. Vaccine has been tested, tracked, and is safe. Make sure everyone understands this.<br><u>**Tweet 2:**</u> But it is misleading to suggest that adverse results in previous mRNA vaccine trials make the Covid 19 vaccines unsafe, particularly when widespread testing of such vaccines has been carried out on humans<br><u>**Tweet 3:**</u> I think the fact that we have been doing vaccine testing for COVID-19 since last March/April has been neglected in communication about vaccine. Virtually everyone I talk to who says "I'll wait to see long term effects" has also had no idea testing began more than a year ago.<br><u>**Tweet 4:**</u> Vaccine development has been much faster for COVID-19. There has been multiple testing all over the world which is why they have been produced quicker.<br><u>**Tweet 5:**</u> The #COVIDVaccines have been tested on a wide range of people to make sure they're safe and effective for everyone.<br><u>**Tweet 6:**</u> The Covid-19 vaccine has been rigorously tested to ensure it meets the highest standards of safety and effectiveness. Please be aware that vaccine misinformation can be spread online. Make sure the information you read is from a reliable source.<br>**Framing A. Scientists have been working on Coronavirus vaccines for decades. The COVID-19 vaccine has been tested and it is safe.** |
| <u>**Tweets DOUBTING the question predication:**</u><br><u>**Tweet 7:**</u> I am seeing folks hesitating to get the COVID-19 vaccine because "it hasn't been adequately tested", but then I see articles about the lingering effects of having COVID-19, and think, "Huh, I'll take my chances with the vaccine."<br>**Framing B. Even if vaccine was not tested for a long time, it is not worth having the lingering effects of COVID-19.** |
| <u>**Tweets ACCEPTING the question predication**</u><br><u>**Tweet 8:**</u> THE COVID-19 VACCINE IS UNTESTED. IT TAKES AT LEAST 5 YEARS TO TEST A VACCINE THEY SAY. THE PROBLEM ROBERT KENNEDY JR HAS WITH THEM IS THEY DONT DO WHAT THEY ARE SUPPOSE TO DO. IT HAS ONLY BEEN 2 YEARS SINCE COVID 19.<br><u>**Tweet 9:**</u> The Covid Vaccines have NOT been adequately tested. Therefore, this vaccine satisfies The Nuremberg Code conditions and definitions for "medical research and experimentation". Coerced Vaccinations would violate The Nuremberg Code of The Geneva Convention.<br>**Framing C. The COVID-19 vaccines have not been tested for at least 5 years, as they should.** |

To infer the framings, we were inspired by work in query-based summarization (Baumel et al., 2016; Yulianti et al., 2018), in which an abstractive summary is created to highlight the most informative aspects of multiple documents that answer a query. In our case, the tweets had the role of documents, and considering that the tweets were already retrieved based on the processing of attitude-evoking questions, two computational linguistics experts selected the discourse units that are shared by a set of tweets, from which the framing was generated, informed by the pyramid method. The pyramid method (Nenkova & Passonneau, 2004) is an empirically grounded method for content selection that quantifies the centrality of viewpoints.

For example, as illustrated in Table 2, in generating the Framing A, first, a discourse unit from Tweet 1 was selected, while a second discourse unit was selected from Tweet 5, to generate the abstraction of the framing, similarly to an abstractive summary. The linguists also have inspected all other discourse units of the tweets, deciding whether they are (a) central to the issues discussed across all tweets sharing the same attitude and (b) offering the same response to the inquiry question before selecting them to be used in the framing. When a discourse unit expressed new content, it was selected for the generation of the framing.

The largest number of framings pertaining to confidence in the COVID-19 vaccines were obtained as answers to the more general question: *Q1(C): How confident are you in the safety of the COVID-19 vaccines*? producing 18 different framings. The smallest number of framings per question were inferred when answering the question asserting the predication that “*Getting vaccinated helps protect those who are unable to be vaccinated against the disease*”, which yielded only one framing: *Getting the COVID-19 vaccine will protect my patients, family and friends who cannot get the jab*. All other questions led to the inference of between 2 and 13 framings per question. Similarly, the largest number of framings pertaining to confidence in the HPV vaccine was obtained in response to the question *Q1(H): How confident are you in the safety of the HPV vaccine*? resulting in 10 different framings. Only one framing was inferred in response to the question asserting the predication that “*Improved living standards, not vaccination, have reduced infectious diseases*”. The framing is: *Vaccination is the most effective way of preventing infectious diseases*. All other questions about confidence in the HPV vaccine led to the inference of between 2 and 8 framings per question.

More interesting was the result of the deeper analysis of the content of the framings, distinguishing between framings that are (a) expressing misinformation; (b) building or eroding the trust in vaccines; (c) exposing possession or lack of vaccine literacy; (d) alluding to morality or (e) articulating civil rights concerns.

### Generating a Taxonomy of Misinformation about Vaccines

The decision of whether a framing contained misinformation was based on finding evidence on the Web, as retrieved by search engines, that the framing expressed known misconceptions, or conspiracy theories. In addition, whenever flawed reasoning was observed, the framing was categorized as misinformation. One researcher with expertise in Web search and an expert on Public Health independently judged the framings that contain misinformation. The two researchers adjudicated their differences and decided that out of the 64 framings inferred for the confidence in the HPV vaccines, 21 of them (33%) expressed misinformation. Similarly, out of the 113 framings inferred for the safety of COVID-19 vaccines, misinformation was present in 38 of them (34%). Table 3 illustrates some examples of misinformation.

**Table 3:**
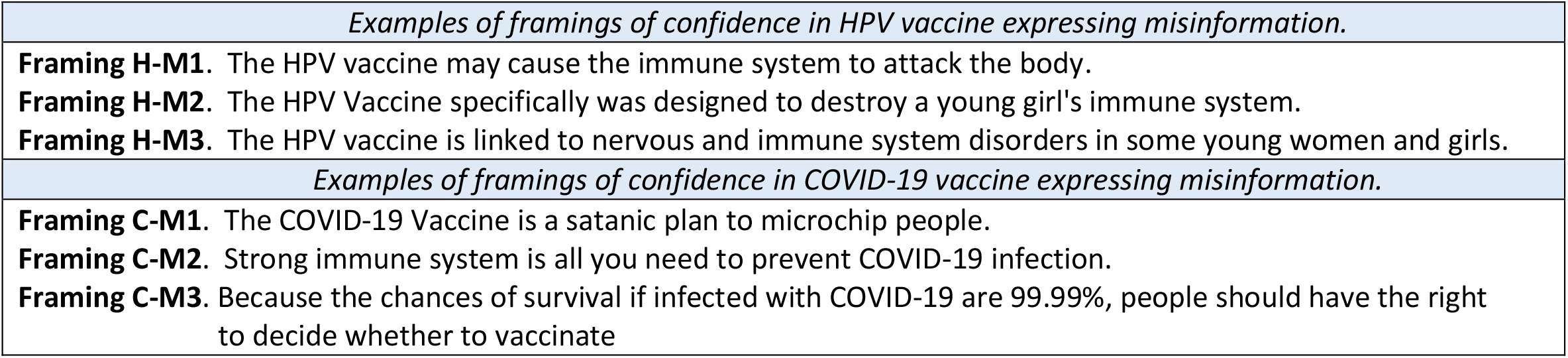
Examples of misinformation expressed in framings inferred from vaccine confidence.

Examples H-M1 to H-M3, pertaining to the HPV vaccine, articulate misinformation about the effects of the vaccine on the immune system. Each framing articulates a different nuance of these effects: H-M1 alerts about the possibility of the immune system to attack the body, H-M2 is packaging the destruction of the immune system as a nefarious design, while H-M3 links not only the disorders of the immune system to the vaccine, but also the disorders of the nervous system. However, all three framings share the theme of the vaccine’s effects on the immune system, an observation that motivated us to derive a taxonomy of misinformation based on the themes and the concerns raised by the misinformation articulated throughout framings. Similarly, the framings C-M1 to C-M3 cover the theme that the COVID-19 vaccines are unnecessary.

All framings inferred for the HPV vaccine that were judged as expressing misinformation were ontologically examined with the goal of discovering common themes and concerns. As in any taxonomy, all framings that shared the same theme were further categorized to uncover the concerns that distinguish framings within the theme. In this way, the taxonomy of misinformation about the HPV vaccine had three layers: (1) themes; (2) concerns within each theme; and (3) framings expressing misinformation. The same methodology of creating a 3-layers taxonomy from the misinformation framings inferred about the confidence in the COVID-19 vaccines generated a second misinformation taxonomy, specific to the COVID-19 vaccine.

### Generating Taxonomies of Trust in Vaccines

While expressing misinformation is also seeding mistrust in the vaccines, to our surprise, many other framings addressed the issue of trust in the safety of vaccines, although not expressing any misinformation. As with judging misinformation, two researchers (one public health expert and a sociolinguist expert) made independent judgements about whether a framing is eroding or increasing trust in vaccine safety, or does not convey any trust issue. The inter-judge agreement was computed using the Kappa score, yielding a score of 0.8 for trust expressed about the HPV vaccine and 0.82 for trust expressed about the COVID-19 vaccines. After adjudicating the judgements, we found that there were 21 framings that increase trust in the safety and 20 frames that erode this trust of the HPV vaccine. Similarly, 27 framings were found to increase the trust while 25 framings eroded trust in the safety of COVID-19 vaccines.

Table 4 illustrates examples of both forms of trust in the HPV or the COVID-19 vaccines. The framings HT-1 to HT-3 illustrated in Table 4 demotivate people from taking the HPV vaccine, by eroding their trust in it, while framings HT+1 to HT+3 do the opposite, motivating people for vaccinating their children against HPV. Framings CT-1 to CT-3 erode the trust in the effects of the COVID-19 vaccine. Interestingly, the framing CT-3, although not directly expressing any misinformation, it refers to a conspiracy theory that widely circulated on Facebook, falsely stating that the French virologist Luc Montagnier, a Nobel prize winner, declared that all those that were vaccinated with the mRNA COVID-19 vaccines will die withing two years. In contrast, the framings CT+1 to CT+3 increase the trust in the COVID-19 vaccines by sharing the theme that they are safe and worth taking. These common themes shared by the framings indicate that two separate taxonomies for trust in the vaccines should be derived, a taxonomy of eroding trust and a taxonomy of building trust in the vaccines. Furthermore, we coded all the framings from *framings*^*HPV*^ and *framings*^*COVID*-19^ that displayed vaccine literacy or lack of. For example, framings HT+1 and HT+2 illustrated in Table 4 were coded as showcasing vaccine literacy, whereas framings HT-1 and HT-2 display lack of vaccine literacy. We found that 17 of the framings used for the HPV vaccine showcased vaccine literacy, while 5 displayed a lack of literacy. Similarly, 27 of the framings used for the COVID-19 vaccine showcased vaccine literacy, while 1 displayed a lack of literacy.

**Table 4:**
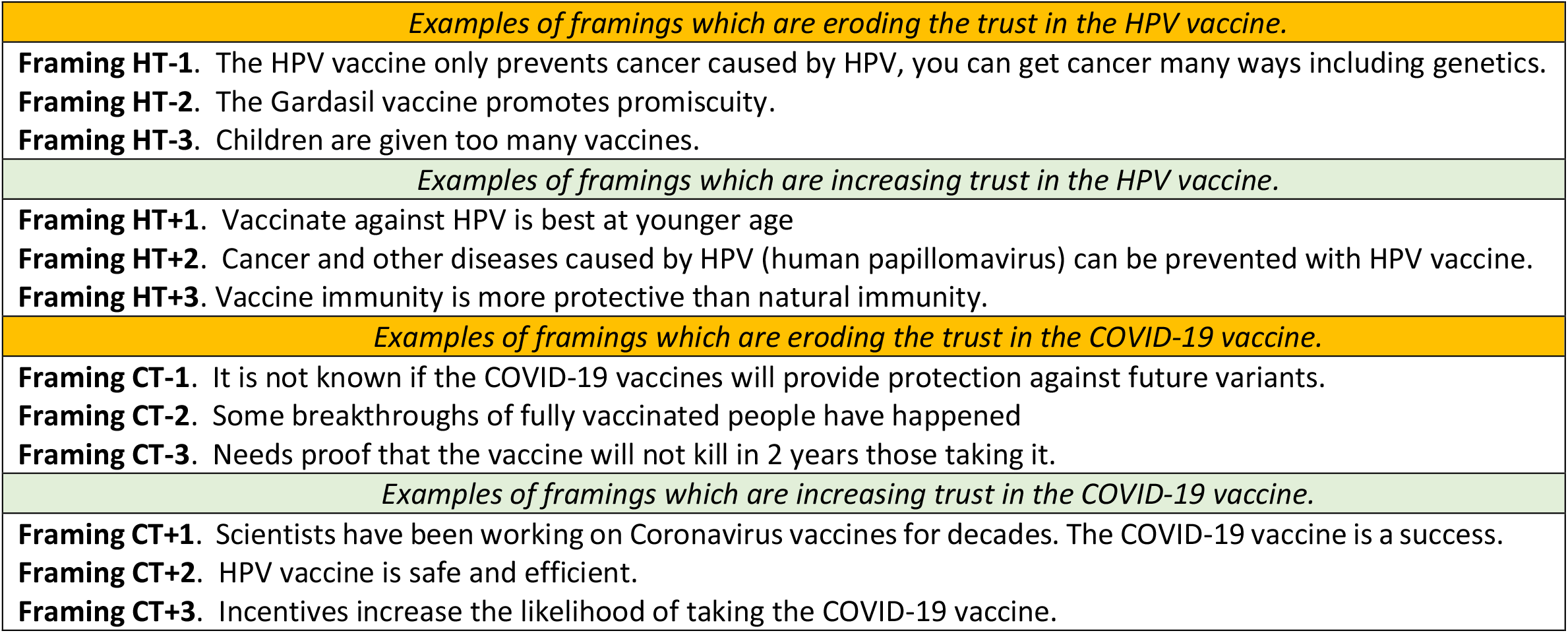
Examples of framings that erode or increase the trust in vaccine confidence.

### Additional Framing Categories

In addition to framings that addressed misinformation or trust in vaccines, a smaller number of framings have addressed civil rights issues. From all the framings that were inferred for the HPV vaccine, 12 framings address civil right issues, while from all the framings inferred for the COVID-19 vaccine 28 framings addressed civil rights issues. Examples of framings that were categorized as expressing civil rights are listed in Table 5.

**Table 5:** Examples of framings that address civil rights or morality.

|  |
| --- |
| *Examples of framings for the HPV vaccine which address civil rights.* |
| **Framing H.CR.1.** Religious exemptions for the Gardasil vaccine guarantee Freedom of Religion. |
| **Framing H.CR.2.** Teenagers should be able to make their own decision about HPV vaccination. |
| **Framing H.CR.3.** It is a parent right to decide if children get vaccinated . |
| *Examples of framings for the COVID-19 vaccine which address civil rights.* |
| **Framing C.CR.1.** COVID-19 vaccine passports are acceptable only for travel but not for other activities. |
| **Framing C.CR.2.** Refusing the COVID-19 vaccine when working in healthcare is unacceptable. |
| **Framing C.CR.3.** Vaccination against COVID-19 should be mandatory. |
| *Examples of framings for the HPV vaccine expressing moral issues.* |
| **Framing H.M.1.** The Gardasil vaccine promotes promiscuity. |
| *Examples of framings for the COVID-19 vaccine expressing moral issues.* |
| **Framing C.M.1.** People that take the COVID-19 vaccine do so out of self-interest, not because of altruism, and thus have no right to morally judge those that refuse the vaccine. |
| **Framing C.M.2.** Patents for the COVID-19 vaccines should be waived, pharmaceutical companies oppose this. |
| **Framing C.M.3.** Pharmaceutical companies test their COVID-19 vaccines in countries with low-cost testing and then sell their vaccines for profit to rich countries. |

We further categorized the framings addressing civil rights in two possible ways: (1) framings implying that vaccination should be prioritized over civil rights (e.g., framing C.CR.3 from Table 5); and (2) framings implying that civil rights should always be prioritized (e.g., framing H.CR.3 from Table 5). Finally, very few framings were coded as expressing morality. Among the framings inferred for the HPV vaccine, only one framing was categorized as expressing morality issues, while for the framings that were inferred for the COVID-19 vaccine, only four framings were categorized as expressing morality issues. Examples of such framings are provided in Table 5.

### Moral Foundations of Framings Concerning Vaccine Confidence

While some of the framings from *framings*^*HPV*^ and *framings*^*COVID*-19^ were categorized as expressing morality issues, we were also interested to explore if implicit moral dimensions could be attributed to all framings. Previous work (Johnson & Goldwasser, 2018, 2019) has shown that there are correlations between stances towards framings and moral convictions that justify the stances. To further explore how the framings of attitudes towards HPV or COVID-19 vaccination, their stances and the moral convictions inform profiles of hesitancy, we considered the Moral Foundations Theory (MFT) (Haidt & Graham, 2007; Haidt & Joseph, 2004), which provides a theoretical framework suggesting that there are five basic moral values which underlie human moral perspectives, emerging from evolutionary, social, and cultural origins. These are referred to as the moral foundations (MF) and include Care/Harm, Fairness/Cheating, Loyalty/Betrayal, Authority/Subversion, and Purity/ Degradation. Table 6 provides the explanations of the MFs.

**Table 6:** Definitions of Moral Foundations from the Moral Foundations Theory.

|  |
| --- |
| **1. Care/Harm:** Care for others, generosity, compassion, ability to feel pain of others, sensitivity to suffering of others, prohibiting actions that harm others. |
| **2. Fairness/Cheating:** Fairness, justice, reciprocity, reciprocal altruism, rights, autonomy, equality, proportionality, prohibiting cheating. |
| **3. Loyalty/Betrayal:** Group affiliation and solidarity, virtues of patriotism, self-sacrifice for the group, prohibiting betrayal of one's group. |
| **4. Authority/Subversion:** Fulfilling social roles, submitting to authority, respect for social hierarchy/traditions, leadership, prohibiting rebellion against authority. |
| **5. Purity/Degradation:** Associations with the sacred and holy, disgust, contamination, religious notions which guide how to live, prohibiting violating the sacred. |
| **6. Non-moral:** Does not fall under any other foundations |

A computational linguist and an expert in public health have independently assigned MFs to all the framings that were inferred for the HPV and the COVID-19 vaccines. The inter-judge agreement was computed using the Kappa score, yielding a score of 0.89 for the HPV vaccine and 0.85 for the COVID-19 vaccines.

### Automatic Discovery of Framings of Vaccine Confidence at Scale

The discovery of tweets from the collection 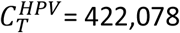 unique tweets that implicitly or explicitly refer to any of the framings from *framings*^*HPV*^ and the discovery of tweets from the collection 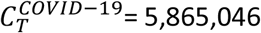 unique tweets that are implicitly or explicitly referring to any of the framings from *framings*^*COVID*-19^ was performed in three steps. First, each framing was used as a query, to retrieve tweets that are deemed relevant to the framing. Second, the tweets were judged by three language experts as being relevant or irrelevant, with inter-judge agreement computed using the Kappa score, yielding a score of 0.82 for relevant tweets for the HPV vaccine framings and 0.84 for relevant tweets for the COVID-19 vaccine framings. Because the third step relied on a Natural Language Processing (NLP) method that is enabled by deep learning, we divided the judged tweets into a training set, a validation set, and a testing set. For the HPV vaccine, the training set contains 4128 tweets, out of which 3703 tweets were judged as referring to a framing. For the same vaccine, the validation set had 459 tweets, out of which 424 tweets referred to a framing, while the testing set had 1147 tweets, out of which 1024 referred to a framing. For the COVID-19 vaccines, the training set contains 7604 tweets, out of which 6684 tweets were judged as referring to a framing, the validation set had 845 tweets, out of which 748 tweets referred to a framing and the testing set had 2113 tweets, out of which 1838 referred to a framing.

Third, we took advantage of an NLP method capable to produce a representation of a Vaccine Attitude Framings Knowledge Graph (VAF-KG) based on the tweets that were judged to explicitly or implicitly convey the framings of vaccine confidence that we discovered. The NLP method was detailed in (Weinzierl & Harabagiu, 2021). This method allowed us to discover that there were 282,651 tweets in 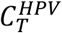 that referred to any of the framings from *framings*^*HPV*^and 1,256,369 tweets in 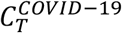 that referred to any of the framings from *framings*^*COVID*-19^.All tweets from the training set that were referring to the same framing were represented by a fully connected graph (FCG), which was further bootstrapped by predicting links from all the tweets available in 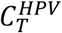, and 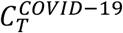 collections which were not yet judged. The prediction was performed using deep learning techniques, detailed in (Weinzierl & Harabagiu, 2021). Essentially, knowledge embeddings are learned for all nodes and edges in the VAF-KG, based on the judgements produced by the language experts. Then scoring functions available from knowledge embedding models provide the estimation of the likelihood of a non-judged tweet to use the same framing as the one represented in one of the FCGs of the VAF-KG. This allowed us to discover 282,651 tweets referring to one or more framings from *framings*^*HPV*^ and 1,256,369 tweets referring to one or more framings from *framings*^*COVID*-19^. Moreover, we discovered that 138,779 Twitter users authored tweets that tweets that implicitly or explicitly refer to any of the framings from *framings*^*HPV*^ and 665,798 Twitter users authored tweets that tweets that implicitly or explicitly refer to any of the framings from *framings*^*COVID*-19^.

### Automatic Recognition of the Stance used in Tweets evoking Framings of Vaccine Confidence

While it is essential to scale up the discovery of the framings in the entire 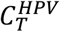 and 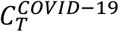 collections, it is equally important to automatically discern the stance the author of the tweet has towards the referred framing, namely they support or reject the framing. To discover the stance of each tweet addressing any of the framings, we relied on a second NLP method using deep learning, detailed in (Weinzierl et al., 2021), by stacking several layers of lexico-syntactic, semantic, and emotion Graph Attention Networks (GATs) (Velickovic et al., 2018) to learn and refine all the possible interactions between these different linguistic phenomena, before classifying a tweet as (a) agreeing; (b) disagreeing or (c) having no stance towards the framing of interest. Stance discovery was made possible by the stance judgements produced by the three language experts which judged whether tweets from the 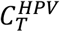 or 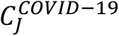 collection are referencing framings from *framings*^*HPV*^ or *framings*^*COVID*-19^, which allowed us to train and test the automatic stance detector, and then use it on the entire 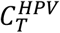 and 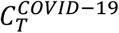 tweet collections. In this way, for each framing it used, each tweet from these collections received a probabilistic distribution [*p*^*Accept*^, *p*^*Reject*^, *p*^*No Stance*^] with respect to the framing. Using the method detailed in (Weinzierl et al., 2021), we discovered that there were 137,261 tweets that accepted some framing from *framings*^*HPV*^ and 54,946 tweets that rejected some framing from *framings*^*HPV*^. Similarly, we have found that there are 877,481 tweets that accepted some framing from *framings*^*COVID*-19^ and 447,716 tweets that rejected some framing from *framings*^*COVID*-19^.

### Discerning Hesitancy Profiles

The recognition of the hesitancy profiles for each vaccine was performed in two steps. First, a Vector User Representation (VUR) was created for any user that authored a tweet from the 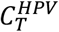 or the 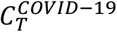 collection. As shown in Figure 2, the VUR has entries for (a) each of the themes from the misinformation taxonomy; (b) each of the themes from the taxonomy for building trust and each of the themes from the taxonomy for eroding trust; (c) a quantification of the vaccine literacy or lack of; (d) a quantification of the impact of civil rights on vaccination; and (e) a quantification of each of the moral foundations. All the values in the VUR are initialized to 0. These values are updated after computing (i) values *v*_*Theme*_ quantifying the conceptualization of misinformation or trust taxonomy themes in each user’s tweets; (ii) values 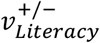 quantifying the vaccine literacy (or lack of) of the framings referred by users in their tweets; (iii) values 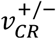 quantifying a user’s preference of vaccination mandates over civil rights (+) or the respect of civil rights, regardless of public health circumstances (-); and (iv) values 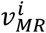 quantifying the support of each of the *i* = 1, …, 9 moral foundations. Central to the computation of these four types of values is the quantification of each framing *Framing*_*X*_ that a user is referring to in its tweets, in a value 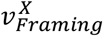.

**Figure 2:**
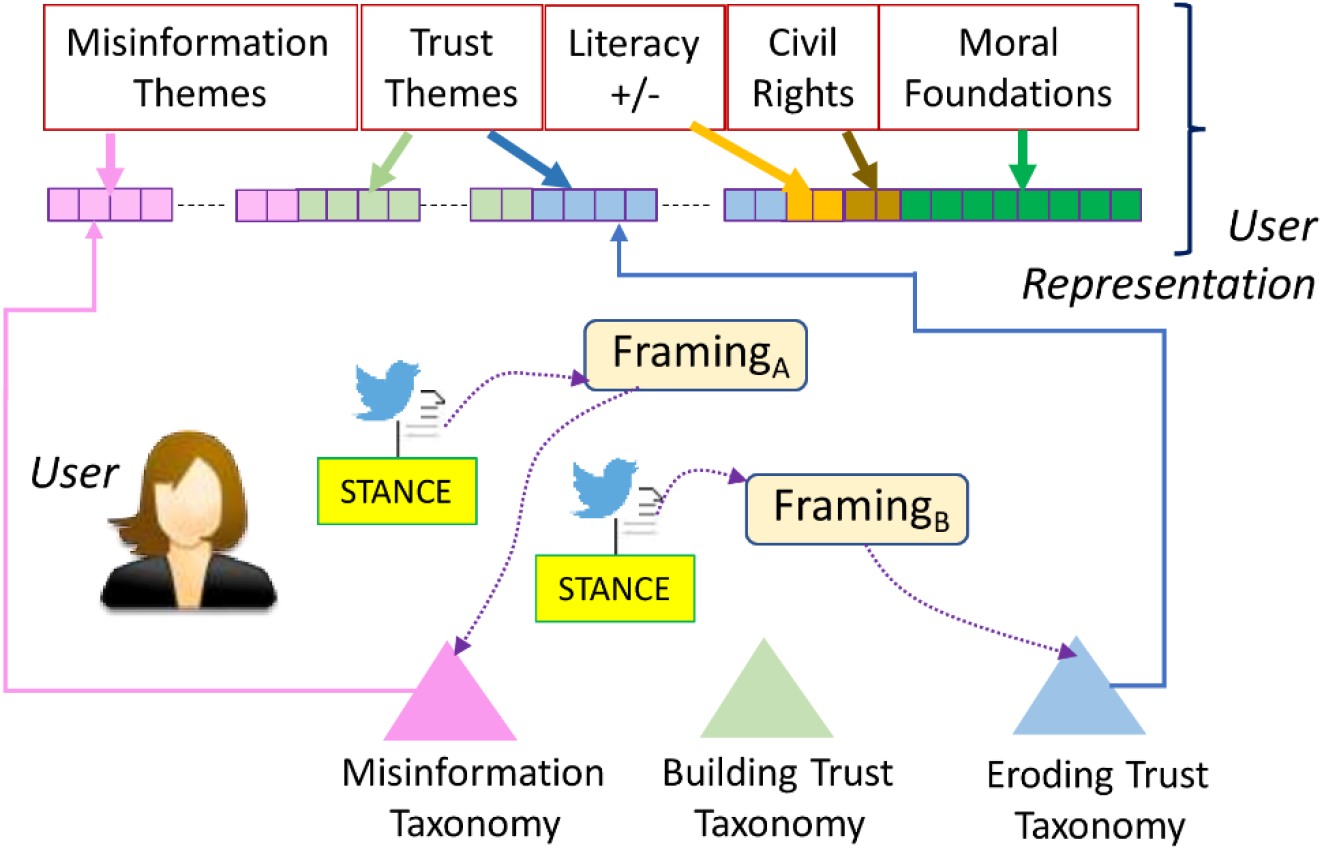
User representation for Profile Discovery.

To compute 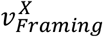 we need to note that any tweet *t* authored by the same user is having in addition to (1) a link to some *Framing*_*X*_, generated by the automatic detection of the framings; (2) a stance that reflects if the tweet *t* is (a) accepting the *Framing*_*X*_, quantified by the probability 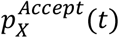; or it rejects the *Framing*_*X*_, quantified by the probability 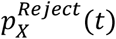; or the tweet has no stance towards *Framing*_*X*_, quantified by the probability 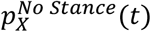, where the distribution 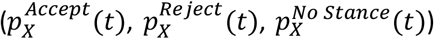 is produced by the automatic stance detection. We decided to make the assignment of 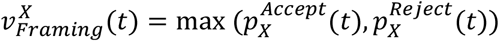, preferring a quantification provided by the dominant stance. Moreover, when the dominant stance was the rejection of the *Framing*_*X*_, we changed the polarity, i.e. 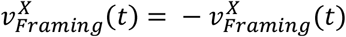, such that all positive values assigned to *Framing*_*X*_ can be interpreted as acceptance of the framing, whereas all negative values can be interpreted as rejection of the framing. All computed 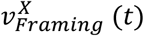 will be withing the interval [−1, +1].

The evaluation of each *v*_*Theme*_ is based on the observation that framings that belong to the misinformation or trust taxonomies can be ontologically characterized by some *Theme*_*Y*_ from one of these taxonomies illustrated in Tables 6, 7, 8 or 9. If a user generates only one tweet *t* that refers to *Framing*_*X*_, then 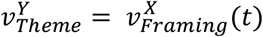. However, when a user generates multiple tweets *t*_*i*_ that referred to the same *Framing*_*X*_, then 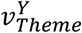 is computed as the average of 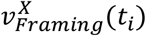. Moreover, if the same user generates tweets which refer to multiple framings *Framing*_*j*_ categorized under the *Theme*_*Y*_ of one of the taxonomies, the value 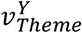 becomes the sum of framing values, i.e.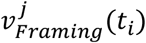. Because of this, for some themes the values in the user representation may be outside the interval [−1,+1].

**Table 7:** A Taxonomy of Misinformation about Confidence in the HPV vaccine.

| **THEME 1: Unsafe HPV vaccine** | **THEME 2: HPV Vaccine Ingredients** |
| --- | --- |
| CONCERN1: Gardasil creator says the vaccine is as dangerous as HPV<br>*CONCERN2: Some scientists explain why the HPV vaccine is unsafe<br>CONCERN3: Multiple countries ban the HPV vaccine<br>CONCERN4: HPV vaccine proven unsafe in Europe<br>CONCERN5: Lack of research about HPV vaccine safety | *CONCERN1: The HPV vaccine contains toxins<br>CONCERN2: The HPV vaccine contains Borax<br>CONCERN3: The HPV vaccine contains lab-engineered DNA |
| **THEME 3: Testing of the HPV Vaccine** | **THEME 4: Alternatives to HPV Vaccine** |
| CONCERN1: HPV vaccine was not tested on boys<br>CONCERN2: Vaccine tested against placebo with high amounts of aluminum<br>CONCERN3: Not tested for carcinogenicity or impairment of fertility<br>CONCERN4: Tested on minorities and underserved because it contains sterility formula | *CONCERN1: Homeopathic medicines are alternatives to HPV vaccine<br>*CONCERN2: Vitamins are alternatives to HPV vaccine<br>CONCERN3: Mushroom extract is an alternative to HPV vaccine |
| **THEME 5: Unnecessary HPV vaccine** | **THEME 6: Effect on Immune System** |
| CONCERN1: The HPV vaccine prevents the same cancer preventable by a Pap smear<br>*CONCERN2: A strong immune system fights off most strains of HPV<br>CONCERN3: Improved living standards, not vaccinations, have reduced infections | CONCERN1: HPV vaccine alters children immune system, even of an unborn child<br>*CONCERN2: HPV vaccine may cause immune system to attack body<br>CONCERN3: HPV vaccine was designed to destroy young girls' immune system |
| **THEME 7: Not effective HPV vaccine** | **THEME 8: Adverse Events of HPV vaccine** |
| CONCERN1: The HPV vaccine causes cancer.<br>CONCERN2: The HPV vaccine does not cover all cancer-causing HPV strains.<br>CONCERN3: The HPV vaccine causes more cancer cases than it prevents. | CONCERN1: Deaths/ Injuries caused by HPV vaccine<br>CONCERN2: The HPV Vaccine causes Infertility<br>CONCERN3: HPV vaccine leads to mental retardation<br>CONCERN4: HPV vaccine causes paralysis |
| **THEME 9: HPV vaccine and promiscuity** | **THEME 10: HPV vaccine information is concealed** |
| CONCERN1: Girls receiving the HPV vaccine will become promiscuous<br>CONCERN2: Not getting the HPV vaccine is not putting others at risk if you are not promiscuous. | CONCERN1: Pharmaceutical companies conceal information about HPV vaccine safety.<br>CONCERN2: Pharmaceutical companies hide cancer cure and release HPV vaccines.<br>*CONCERN3: The Government conceals information about the safety of HPV vaccines. |

**Table 8:** A Taxonomy of Misinformation about Confidence in the COVID-19 Vaccines.

| **THEME 1: Unsafe COVID-19 vaccine** | **THEME 2: COVID-19 Vaccine Ingredients** |
| --- | --- |
| CONCERN1: Vaccine unsafe because the virus is a bioweapon.<br>*CONCERN2: Some scientists explain why the COVID-19 vaccine is unsafe<br>CONCERN3: Bill Gates admits the COVID-19 vaccine is unsafe<br>CONCERN4: The COVID-19 vaccine makes you gay<br>CONCERN5: The COVID-19 vaccine makes you 5G compatible<br>CONCERN 6: The COVID-19 vaccine renders pregnancies risky | *CONCERN1: The COVID-19 vaccine injects a toxin in your bloodstream<br>CONCERN2: The COVID-19 vaccine uses nanotechnology<br>CONCERN3: The vaccine is gene therapy that activates a toxin in your body<br>CONCERN4: The mRNA vaccine contains the virus |
| **THEME 3: Testing of the COVID-19 Vaccine** | **THEME 4: Alternatives to COVID-19 Vaccine** |
| CONCERN1: No-long term studies of side-effects of COVID-19 vaccine<br>CONCERN2: No COVID-19 vaccine efficacy or safety data<br>CONCERN3: Vaccine has not been tested for at least 5 years | *CONCERN1: Homeopathic medicines are alternatives to HPV vaccine<br>*CONCERN2: Vitamins are alternatives to COVID-19 vaccine<br>CONCERN3: Hydroxychloroquine as alternative to vaccine<br>CONCERN4: Garlic as alternative to vaccine<br>CONCERN5: Mushroom extract is an alternative to HPV vaccine |
| **THEME 5: Unnecessary COVID-19 vaccine** | **THEME 6: Effect on Immune System** |
| CONCERN1: The vaccine is a satanic plan to microchip population<br>*CONCERN2: A strong immune system is all you need.<br>CONCERN3: Chances of surviving infection are 99.99%<br>CONCERN4: People with severe allergies should not be vaccinated | CONCERN1: COVID-19 vaccine overwhelms the immune system<br>*CONCERN2: COVID-19 vaccine may cause immune system to attack body<br>CONCERN3: COVID-19 vaccine overrides the immune system |
| **THEME 7: Not effective COVID-19 vaccine** | **THEME 8: Adverse Events of COVID-19 vaccine** |
| CONCERN1: The vaccine does not protect against COVID-19 infection<br>CONCERN2: Natural Immunity lasts longer than vaccine-induced immunity.<br>CONCERN3: People better protected by immunity gained through infection then immunity gained through vaccination | CONCERN1: COVID-19 vaccine interacts with people's DNA<br>CONCERN2: COVID-19 vaccine replaces the genetic code with a synthetic one |
|  | **THEME 10: COVID-19 vaccine information is concealed** |
| . | CONCERN1: Pharmaceutical companies conceal information about breakthroughs and reinfections.<br>CONCERN2: The Federal Government lied about vaccines to reduce the information about COVID-19 treatments.<br>*CONCERN3: The Government conceals information about the safety of COVID-19 vaccines. |

The evaluation of the values 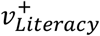 result from taking the average value of all 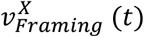 that were annotated as exhibiting vaccine literacy, while 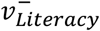 results from taking the average value of all 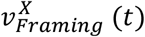 that showcase lack of vaccine literacy, given every tweet *t* of a user. Finally, the evaluation of the values 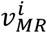, for each of the *i* = 1, …, 9 moral foundations, listed in Table 10, is generated by taking the average value of all 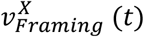, for each framing that was annotated with the moral value *i*. When all users VUR were generated, we were able to discern the user profiles, by using the k-Means clustering algorithm (Lloyd, 1982), experimenting with *K* = 2, …, 10 possible clusters. The final number of clusters was determined by the Elbow method [Thorndike 1953]. We found *K* = 5 to be optimal for both HPV and COVID-19 vaccine hesitancy profiles.

**Table 9:** The Taxonomy for building Trust in HPV vaccines and the Taxonomy for eroding Trust in the HPV vaccines.

| Taxonomy of Building TRUST in HPV Vaccines | Taxonomy of Eroding TRUST in HPV Vaccines |
| --- | --- |
| **<u>THEME 1: Trust in Safety of HPV vaccines</u>**<br>*CONCERN 1: HPV vaccine is safe; I gave it to my kids!<br>*CONCERN 2: Information about the safety of vaccines is available on VAERS.<br>CONCERN 3: There are no serious allergy reactions to HPV reported.<br>**<u>THEME 2: Motivation for taking HPV vaccines</u>**<br>CONCERN 1: Cancer and other diseases caused by HPV (human papillomavirus) can be prevented with HPV vaccine.<br>CONCERN 2: Vaccinate against HPV is best at younger age<br>CONCERN 3: Vaccines can save lives and eliminate diseases.<br>CONCERN 4: Vaccination is the most effective way of preventing infectious diseases.<br>*CONCERN 5: Vaccine immunity is more protective than natural immunity.<br>**<u>THEME 3: Trust in Effects of HPV vaccines</u>**<br>CONCERN 1: HPV vaccine improves children's immune systems.<br>CONCERN 2: The HPV vaccine does not suppress the immune system like the natural HPV infection.<br>CONCERN 3: Not having side effects after the HPV vaccine depends on the immune system.<br>CONCERN 4: HPV vaccine cannot cause cancer or any other disease.<br>**<u>THEME 4: Trust in role of HPV vaccines for Public Health</u>**<br>CONCERN 1: HPV vaccine can completely eradicate cervical cancer.<br>CONCERN 2: Herd Protection through vaccination.<br>CONCERN 3: Vaccinations are the key to controlling infectious disease and the secondary conditions associated with infection<br>CONCERN 4: The benefits of the HPV vaccine extend to people who aren't vaccinated — meaning the more people who are vaccinated, the better.<br>CONCERN 5: To eradicate several cancers, HPV vaccination should be compulsory and free.<br>**<u>THEME 5: Trust in the role of pharmaceutical companies for preventing cancer with HPV vaccines</u>**<br>*CONCERN 1: Vaccines are not profitable for pharmaceutical companies<br>CONCERN 2: HPV Vaccines prevent cancer, regardless of the profit of pharmaceutical companies.<br>**<u>THEME 6: Trust in Doctors/ Science</u>**<br>CONCERN 1: Safety and efficacy of HPV vaccines supported/explained by doctors.<br>CONCERN 2: Safety and efficacy of HPV vaccines supported/explained by science. | **<u>THEME 1: Lack of Trust in Safety of HPV vaccines</u>**<br>CONCERN 1: HPV vaccine may provoke autoimmune diseases.<br>CONCERN 2: Children/Teens with allergy should not be vaccinated.<br>CONCERN 3: Vaccines alter the immune system of a child.<br>**<u>THEME 2: De-Motivation in taking the HPV vaccines</u>**<br>CONCERN 1: The HPV vaccine only prevents cancer caused by HPV; you can get cancer many ways including genetics.<br>CONCERN 2: Children are given too many vaccines<br>CONCERN 3: The Gardasil vaccine promotes promiscuity.<br>**<u>THEME 3: Lack of Trust in Effects of HPV vaccines</u>**<br>CONCERN 1: There are some side effects specific to age, but they aren't really about the child's immune system.<br>**<u>THEME 4: Lack of Trust in role of HPV vaccines for Public Health</u>**<br>CONCERN 1: Why mandate a children vaccine that protects against cancer that appears later in life.<br>CONCERN 2: If vaccines work, the vaccinated have nothing to fear from the unvaccinated.<br>**<u>THEME 5: Lack of Trust in the role of pharmaceutical companies for preventing cancer with HPV vaccines</u>**<br>*CONCERN 1: Pharmaceutical companies produce ineffective HPV vaccine for profit<br>CONCERN 2: Mandating the vaccines is benefitting the pharmaceutical companies.<br>**<u>THEME 6: Lack of Confidence in Doctors/ Science</u>**<br>CONCERN 1: Doctors do not tell the truth about HPV vaccines.<br>CONCERN 2: Researchers/scientists do not tell the truth about HPV vaccines<br>CONCERN 3: Casting doubt about published research of HPV vaccine.<br>**<u>THEME 7: Lack of Trust in Ingredients of HPV vaccines</u>**<br>CONCERN 1: HPV Vaccine contains aluminum adjuvant<br>**<u>THEME 8: Lack of Trust in the testing of the HPV vaccine</u>**<br>CONCERN 1: HPV vaccine rushed into use without adequate testing.<br>CONCERN 2: Distrust of FDA testing procedures.<br>CONCERN 3: HPV vaccine was not tested if it causes cancer. |

**Table 10:** The Taxonomy for building Trust in COVID-19 vaccines and the Taxonomy for eroding Trust in the COVID-19 vaccines.

| Taxonomy of Increasing TRUST in COVID-19 Vaccines | Taxonomy of Eroding TRUST in COVID-19 Vaccines |
| --- | --- |
| **<u>THEME 1: Trust in Safety of COVID-19 vaccines</u>**<br>*CONCERN 1. HPV vaccine is safe and efficient. Scientists have been working on Coronavirus vaccines for decades. The COVID-19 vaccine has been tested, tracked and it is safe.<br>*CONCERN 2. The Government has provided plenty of safety information about the COVID-19 vaccines<br>**<u>THEME 2: Motivation for taking the COVID-19 vaccines</u>**<br>CONCERN 1: Given the risks of COVID-19, it is unlikely that building natural immunity is a good idea.<br>CONCERN 2. Even if vaccine not tested for a long time, it is not worth having the lingering effects of COVID-19.<br>CONCERN 3. The COVID-19 vaccine is not making you immune to the infection, but it is mitigating the effect of the infection.<br>CONCERN 4: The vaccines trigger your body to naturally create immunity more reliably than getting COVID.<br>*CONCERN 5. Natural antibodies last a few months and only partially protect you, while the vaccine instructs your immune system to produce antibodies that last longer and protect you better.<br>CONCERN 6: COVID-19 vaccines protect against the emerging variants.<br>CONCERN 7. Incentives for taking the COVID-19 vaccine.<br>CONCERN 8: Children over 12 should be vaccinated to avoid distant learning.<br>**<u>THEME 3: Trust in Effects of COVID-19 vaccines</u>**<br>CONCERN 1: Vaccination against COVID-19 Strengthens the immune system.<br>CONCERN 2: More likely to get thrombosis from flying economy than from Astra Zeneca.<br>CONCERN 3: Johnson and Johnson COVID-19 vaccine allegedly preferred over Pfizer or Moderna for those with allergies.<br>**<u>THEME 4: Trust in role of COVID-19 vaccines for Public Health</u>**<br>CONCERN 1: Getting the COVID-19 vaccine will protect my patients, family and friends who cannot get the jab. Vaccination is key in protecting yourself and others against COVID-19.<br>CONCERN 2: Because of the risk of new variants that may escape the COVID-19 vaccines, the decision to not vaccinate puts everyone at risk.<br>CONCERN 3: People can choose not to vaccinate if they pay a higher medical bill when needing hospitalization because of COVID-19.<br>**<u>THEME 5: Trust in role of pharmaceutical companies in fighting COVID-19 infections</u>**<br>*CONCERN 1: COVID-19 vaccines are not profitable for pharmaceutical companies unless they are safe and effective.<br>**<u>THEME 6: Trust in Ingredients of COVID-19 vaccines</u>**<br>CONCERN 1: The mRNA COVID-19 Vaccine uses the RNA of COVID-19 which leaves your body soon after you get vaccinated.<br>**<u>THEME 7: Trust in Testing of COVID-19 vaccines</u>**<br>CONCERN 1: COVID-19 vaccination trials for children proven vital | **<u>THEME 1: Lack of Trust in Safety of COVID-19 vaccines</u>**<br>CONCERN 1: if the COVID-19 vaccines are completely safe, why no accidental death policy.<br>CONCERN 2: Vaccine exemptions should be available because the COVID-19 vaccines are experimental.<br>CONCERN 3: Children should not be vaccinated against COVID-19 because there is no legal accountability for adverse events.<br>CONCERN 4: Governments of the Western world advocate COVID-19 vaccines without long-term studies of safety or efficacy, while Asian governments control COVID-19 infections with traditional safety measures.<br>**<u>THEME 2: De-Motivation in taking the COVID-19 vaccines</u>**<br>CONCERN 1: The COVID-19 vaccine does not provide immunity against infection.<br>CONCERN 2: People having severe allergy should be monitored for 30 minutes after receiving the Pfizer COVID-19 vaccine.<br>CONCERN 3: It is not known if the COVID-19 vaccines will provide protection against future variants.<br>**<u>THEME 3: Lack of Trust in Effects of COVID-19 vaccines</u>**<br>CONCERN 1: Lack of confidence in mRNA vaccines and their long-term effects.<br>CONCERN 2: Needs proof that the vaccine will not kill in 2 years those taking it.<br>CONCERN 3: Wait one year to see if there are no long-lasting side effects.<br>CONCERN 4: Astra Zeneca vaccine determines blood clots.<br>CONCERN 5: Fear vaccine may worsen existing conditions.<br>**<u>THEME 4: Lack of Trust in role of COVID-19 vaccines for Public Health</u>**<br>CONCERN 1: People that do not believe that COVID-19 is real or do not believe that masks work should not be receiving vaccines be exempt from the vaccination.<br>CONCERN 2: Because the authorities advocate so hard for COVID-19 vaccination should be the main reason for refusing the vaccine.<br>CONCERN 3: Preference for getting COVID-19 and fighting it off than vaccinating.<br>**<u>THEME 5: Lack of Trust in role of pharmaceutical companies in fighting COVID-19 infections</u>**<br>*CONCERN 1: Pharmaceutical companies will profit because COVID-19 waves will never end, thus requiring annual boosters.<br>**<u>THEME 6: Lack of Trust in Ingredients of COVID-19 vaccines</u>**<br>CONCERN 1: The COVID-19 Vaccine injects the dead SARS-COV2 virus in your body<br>**<u>THEME 7: Lack of Trust in Testing of COVID-19 vaccines</u>**<br>CONCERN 1: COVID-19 vaccines were 'rushed,' so they could still be unsafe.<br>CONCERN 2: AstraZeneca used outdated information in its COVID-19 vaccine trials. |

## RESULTS

Figure 3 illustrates the results that enabled us to answer our first research question:

**Figure 3:**
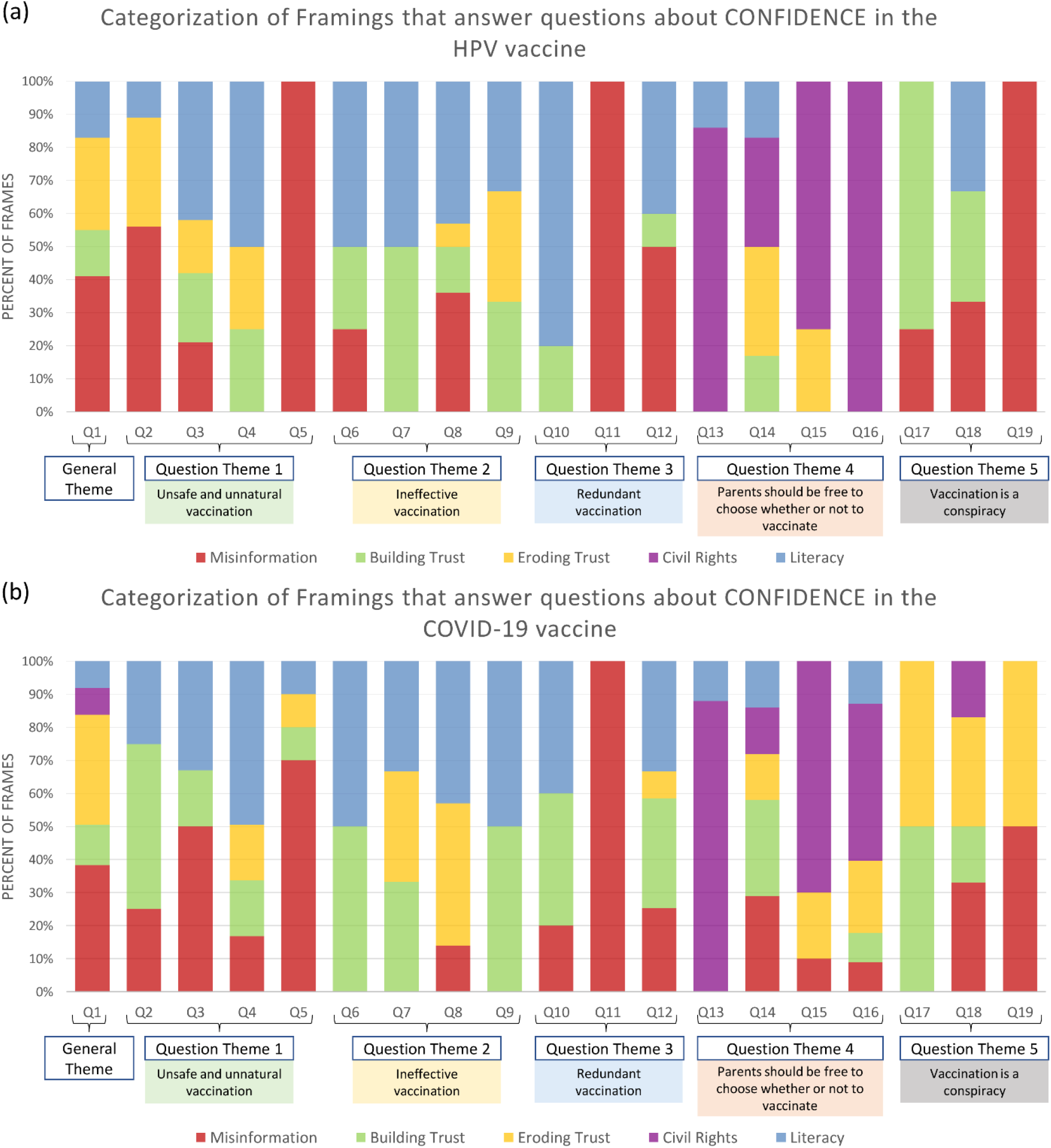
Distribution of Framings Categories. Framings are answers for the questions of varied confidence themes.

### RQ1: How is confidence in the HPV and the COVID-19 vaccines framed in the Twitter discourse?

The Question/Answering (Q/A) framework of this study, illustrated in Figure 1, enables the inference of 64 HPV framings and 113 COVID-19 framings as answers to the general theme question Q1: “*How confident are you in the safety of the HPV/COVID-19 vaccine*? ”, and a set of 18 questions from the Vaccine Confidence Repository.

When the Vaccine Confidence Repository (VCR) for the HPV vaccine was introduced in Rossen et al. (2019), the VCR questions were also grouped into five major question themes, illustrated in Figure 3. We created a similar VCR consisting of questions for the COVID-19 vaccine. The entire list of VCR questions that were used is available in the supplemental material. Figure 3 illustrates the distribution of the framing categories across all question themes. Surprisingly, for the HPV vaccine, misinformation framings were inferred across all question themes, except for question Theme 4, where civil rights dominated. We expected to find a lot of misinformation in the framings answering the questions from theme 5, but we were startled to find plenty of misinformation answering questions from themes 1, 2 and 3. Moreover, the framings answering the general theme also contained misinformation, indicating that misinformation is pervasive in the framing of confidence in the HPV vaccines. As shown in Figure 3, misinformation (indicated in red shading) is also present in framings answering questions about confidence in the COVID-19 vaccines.

We also noticed that there is a higher percentage of framings that erode trust (yellow shading) in the COVID-19 vaccines than in the HPV vaccine. Surprisingly, there is a substantial percentage of framings that build trust across both vaccines. Framings involving civil right issues were inferred as answers mostly to questions from Theme 4 about parents’ right to decide whether to vaccinate their children for both vaccines. Vaccine literacy (blue shading) seems to be present in most framings except Theme 5 (vaccine conspiracy theories) in the case of COVID-19.

When scaling up the discovery of framings at the level of the entire Tweet collection, the system that we used obtained Precision of 80.1%, Recall of 83.2%, and F1 score of 81.6% when evaluated om the test collection for the HPV vaccine and Precision of 71.5%, Recall of 75.8%, and F1 score of 73.6% when evaluated in the test collection for the COVID-19 vaccines. The stance detection system we used recognized the “Accept” stance with an F1 score of 86.5%; the “Reject” stance with F1 score of 70.5% for framings about confidence in the HPV vaccines. The system recognized the “Accept” stance with an F1 score of 87.6% and the “Reject” stance with F1 score of 71.5% when operating on framings of the COVID-19 vaccine. These results indicate that these two automatic systems performed quite well when performing on the entire 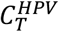 and 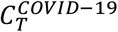 collections.

The Misinformation Taxonomies for each vaccine provide answers to:

### RQ2: What specific misinformation about the HPV and COVID-19 vaccines is propagated on Twitter?

Ten misinformation themes were discovered in the Misinformation Taxonomy for the HPV vaccine, illustrated in Table 7 while nine misinformation themes were discovered for COVID-19 vaccination, illustrated in Table 8. Asterix among concerns denotes that these concerns were common across vaccines. Although higher order themes were similar across vaccines, with the exception of HPV vaccination having one additional promiscuity theme, concerns across vaccines differed in number and content sharing only 21% of concerns. Of the 33 concerns, only 7 were shared across vaccines. This suggests that misinformation is tailored to the worries that are vaccine specific.

Additionally, because misinformation taxonomies have three layers of abstraction namely: (L1)themes →(L2) concerns→(L3)framings→tweets, it is important to also compare the number of framings expressing misinformation in both taxonomies, as it represents he lowest level of abstraction. The number of framings that expressed misinformation is quite different for each vaccine: 21 framings for the HPV vaccine and 38 framings for the COVID-19 vaccine.

Two different Trust taxonomies discovered for each vaccine provided the answers to:

### RQ3: What trust issues are associated with the HPV and COVID-19 vaccines in Twitter conversations?

Table 9 illustrates these taxonomies for the HPV vaccine, whereas Table 10 illustrates both taxonomies for the COVID-19 vaccine. Trust in the HPV vaccine is characterized by 6 themes, concerning (a) vaccine safety; (b) motivation to vaccinate; (c) effect of the vaccine and (d) the role the vaccine plays for public health; (e) trust in the role of pharmaceutical companies for preventing cancer with HPV vaccines and (f) confidence in doctors and/or science. The taxonomy for eroding trust in the HPV vaccine has six themes expressing directly opposed predications to the themes from the taxonomy of building trust, as well as two additional themes, one concerning the lack of trust in the ingredients of the HPV vaccine, the other tackling the lack of trust in testing of the vaccine.

The taxonomy encoding knowledge that builds trust in the HPV vaccines used 21 different concerns spread across 6 different themes, while the taxonomy that encodes knowledge that erodes the trust in the HPV vaccine used 18 concerns distributed across 8 different themes. While 6 out of 8 of the themes of taxonomy that erode trust are antonyms of the themes encoded in taxonomy that building trust in HPV vaccines, contradictory relations between all their concerns cannot be established. For example, Concern 1 of Theme 1 encoded in trust building taxonomy is contradicted by all three concerns of Theme 1 from that trust eroding taxonomy, but Concern 2 of Theme 1 encoded in the trust building taxonomy is not contradicted by any of the concerns of Theme 1 from the opposite taxonomy. This indicates that opposing themes may encode concerns that characterize only one of the taxonomies. Moreover, it shows that some encoded concerns may be contradicted by more than one of the concerns of the opposing theme in the other taxonomy. The interpretation of the structure of the two trust taxonomies listed in Table 9 leads to the conclusion that trust issues associated with the HPV vaccine are not only conceptually diverse, but also characterized by diffused polarization along some of the concerns.

The taxonomies for building and eroding trust in the COVID-19 vaccines were discerned in the same way as were the trust taxonomies encoding trust in the HPV vaccine. As shown in Table 10, the taxonomy encoding the knowledge that builds trust in the COVID-19 vaccines is characterized by seven themes, out of which are five are identical to five themes from the taxonomy that builds trust in the HPV vaccines. All themes from the taxonomy encoding knowledge about building trust in the COVID-19 vaccine have opposite themes in the taxonomy that encodes knowledge about the erosion of trust in the COVID-19. Compared with the themes of the trust taxonomies discerned for the HPV vaccine, the theme concerning the trust in doctors or scientists is missing in the trust taxonomies for the COVID-19 vaccine.

When comparing the taxonomy encoding knowledge that builds trust in the HPV vaccine with the same taxonomy for the COVID-19 vaccine, we observed only 4 common concerns, which are marked with * in Tables 9 and 10. This represents 23% of the trust building concerns encoded for the HPV vaccines; and 21% of the trust building concerns for the COVID-19 vaccine, almost the same percentage of shared concerns as those observed across the misinformation taxonomies. However, the same comparison on the trust eroding taxonomies leads to the observation that here is only one shared concern, marked with * in Tables 9 and 10. This results in 5% of shared concerns for the eroding trust in HPV vaccines and 6% of the shared concerns for the eroding trust in the COVID-19 vaccines.

The annotations of the implied Moral Foundations (MFs) provide the answer the research question:

### RQ4: What moral dimensions characterize the confidence in the HPV and COVID-19 vaccines on Twitter?

Overall, there were 111 moral foundations (MFs) annotated in the 64 confidence framings for the HPV vaccine and 230 MFs annotated in the 113 confidence framings used for the COVID-19 vaccines. Interestingly, 1 framing was coded with 4 MFs, 32 with 3 MFs, 95 with 2 MFs and 48 framings with one MF. The predominant MF in the confidence framings for the HPV vaccine was a tie between Authority and Subversion, while the predominant MF in the framings for the COVID-19 vaccine was Harm.

This indicates that the most common approach by which each vaccine is morally framed in public discourse shifts depending on the vaccine. The framings that expressed misinformation mostly implied the MF Subversion. The framings that conveyed trust erosion in vaccines predominantly conveyed the MFs Subversion and Harm, while the framings that built trust in the vaccine implied the MFs Care and Authority. Framings that involved civil rights issues were predominantly implying the MF Fairness, while framings that involved literacy issues were predominately implying the MF Care.

Finally, the hesitancy profiles that we derived inform the answer to the research question:

### RQ5: What hesitancy profiles can be discerned from Twitter for the HPV and COVID-19 vaccines?

The hesitancy profiles derived for the HPV vaccine are detailed in Table 11, which lists five profiles, along with the number of users in each hesitancy profile, defined by prototypical vector user representations. The interpretation of the *Promoters* of the HPV vaccine, based on their prototypical user representation, is that they are Twitter users who actively promote the HPV vaccine, either because they are public health officials, doctors, scientists or just extremely vaccine literate Twitter users. They are extremely trusting to the point of rejecting the theme of “Lack of Trust in Safety of HPV Vaccines” and actively push mandatory or expanded vaccination programs, as indicated by the high value attributed to the belief (+0.77) attributed to the belief that vaccines are more important than civil rights. These users focus on Morality Foundations of Care, Authority, Loyalty, and Fairness and actively reject Subversion and Harm.

**Table 11:** Twitter Hesitancy Profiles Discovered for the HPV vaccine.

|  |  |
| --- | --- |
| **Profile 1: PROMOTERS** (14,403 users; 21%)<br><u>Themes for Eroding Trust in Vaccine:</u><br>-0.05: Lack of Trust in Safety of HPV Vaccines<br><u>Themes for Building Trust in Vaccine:</u><br>+1.01: Motivation for Taking HPV Vaccines<br>+0.99: Trust in Role of HPV Vaccines for Public Health<br>+0.77: Trust in the Role of Pharmaceutical Companies for preventing Cancer with HPV Vaccines<br>+0.07: Trust in Doctors/Science<br><u>Vaccine Literacy:</u><br>+2.08: Having literacy \| -0.08: Lacking literacy<br><u>Civil Rights:</u><br>+0.77: Vaccines more important than civil rights<br><u>Moral Foundations:</u><br>+2.81: Care \| +2.77: Authority \| +0.89: Loyalty \| +0.89: Fairness \| -0.11: Subversion \| -0.10: Harm \| -0.09: Degradation \| -0.05: Purity | **Profile 2: TRUSTERS** (19,338 users; 28%)<br><u>Themes for Building Trust in Vaccines:</u><br>+0.45: Motivation for Taking HPV Vaccines<br>+0.27: Trust in Role of HPV Vaccines for Public Health<br>+0.17: Trust in Doctors/Science<br>+0.10: Trust in Effects of HPV Vaccines<br>+0.09: Trust in the Safety of HPV Vaccines<br><u>Vaccine Literacy:</u><br>+0.93: Having literacy \| -0.05: Lacking literacy<br><u>Civil Rights:</u><br>+0.06: Civil rights above all<br><u>Moral Foundations:</u><br>+0.94: Care \| +0.84: Authority \| +0.23: Fairness \| +0.19: Loyalty \| +0.11: Betrayal \| -0.11: Subversion<br>-0.08: Harm \| -0.06: Degradation |
| **Profile 3: DEBUNKERS** (21,904 users; 32%)<br><u>Misinformation Themes:</u><br>-0.10: Not effective HPV Vaccine<br>-0.06: Adverse Events of HPV Vaccine<br><u>Themes for Eroding Trust in Vaccine:</u><br>-0.12: Lack of Trust in Safety of HPV Vaccines<br>-0.11: Lack of Trust in the Testing of the HPV Vaccines<br>-0.09: Lack of Trust in Role of HPV Vaccines for Public Health<br>-0.08: De-Motivation in Taking the HPV Vaccines<br><u>Themes for Building Trust in Vaccine:</u><br>-0.05: Trust in the Safety of HPV Vaccines<br><u>Vaccine Literacy:</u><br>-0.14: Having literacy \| -0.25: Lacking literacy<br><u>Civil Rights:</u><br>-0.20: Civil rights above all<br><u>Moral Foundations:</u><br>-0.35: Degradation \| -0.35: Subversion \| -0.32: Authority \| -0.25: Purity \| -0.25: Harm \| -0.18: Care \| -0.13: Cheating \| -0.07: Betrayal | **Profile 4: MISINFORMERS** (4,673 users; 7%)<br><u>Misinformation Themes:</u><br>+0.91: Information about HPV Vaccines is Concealed<br>+0.69: Not effective HPV Vaccine<br>+0.18: Unsafe HPV Vaccine<br>+0.16: HPV Vaccine Ingredients<br>+0.14: Testing of HPV Vaccine<br>+0.07: Adverse Events of HPV Vaccine<br><u>Themes for Eroding Trust in Vaccine:</u><br>+0.38: Lack of Trust in the Role of Pharmaceutical Companies for Preventing Cancer with HPV Vaccines<br>+0.16: Lack of Trust in Doctors/Science<br>+0.13: Lack of Trust in Safety of HPV Vaccines<br>+0.07: De-Motivation in Taking the HPV Vaccines<br>+0.06: Lack of Trust in the Testing of the HPV Vaccines<br><u>Themes for Building Trust in Vaccine:</u><br>+0.70: Trust in the Role of Pharmaceutical Companies for Preventing Cancer with HPV Vaccines<br>-0.09: Trust in the Safety of HPV Vaccines<br>-0.08: Trust in Doctors/Science<br><u>Vaccine Literacy:</u><br>-0.12: Having literacy \| +0.12: Lacking literacy<br><u>Civil Rights:</u><br>+0.09: Civil rights above all<br><u>Moral Foundations:</u><br>+2.80: Betrayal \| +1.59: Harm \| +0.61: Authority \| -0.06: Care<br>+0.58: Cheating \| +0.41: Subversion \| +0.27: Degradation |
| **Profile 5: SKEPTICS** (8,787 users; 13%)<br><u>Misinformation Themes:</u><br>+0.32: Unsafe HPV Vaccine \| +0.21: Not effective HPV Vaccine<br>+0.11: Testing of HPV Vaccine \| +0.07: HPV Vaccine Ingredients<br>+0.11: Information about HPV Vaccines is Concealed<br>+0.09: Adverse Events of HPV Vaccine \|<br><u>Themes for Eroding Trust in Vaccine:</u><br>+0.27: Lack of Trust in Doctors/Science<br>+0.19: Lack of Trust in Safety of HPV Vaccines<br>+0.12: De-Motivation in Taking the HPV Vaccines<br>+0.05: Lack of Trust in the Testing of the HPV Vaccines<br><u>Themes for Building Trust in Vaccine:</u><br>-0.11: Trust in Doctors/Science<br><u>Vaccine Literacy:</u><br>+0.23: Lacking literacy \| -0.09: Having literacy<br><u>Civil Rights:</u><br>+0.18: Civil rights above all<br><u>Moral Foundations:</u><br>+0.61: Subversion \| +0.40: Harm \| +0.35: Betrayal \| +0.24: Cheating \| +0.22: Degradation \| +0.17: Purity \| +0.06: Fairness |  |

The interpretation of the profile of the *Trusters* in the HPV vaccine, based on their prototypical user representation available in Table 11, is that they are Twitter users who generally build trust in the HPV vaccine and are showing literacy towards the vaccine. They widely trust science and doctors, but do not necessarily think the HPV vaccine should be mandatory, while focusing on Care and Authority. Interestingly, the *Debunkers* are users who debunk misinformation and do not believe in the erosion of trust in the HPV vaccine. They do not believe that civil rights should be above all, and they actively reject Degradation, Subversion, Authority, Purity, and Harm. On the opposite end of hesitancy are the *Misinformers*, who actively propagate HPV vaccine misinformation of all kinds, and widely distrust the HPV vaccine. These users lack vaccine literacy and generally oppose vaccine mandates, over civil rights.

They also sow distrust in the vaccine, with a singular exception regarding Pharmaceutical Companies. These users also have significant values for the Moral Foundations of Betrayal and Harm. The *Skeptics* seem to mistrust doctors, science, and the testing of the HPV vaccine. They dip their toes into misinformation, but do not widely accept or promote it. These Twitter users present some vaccine literacy, but much less than the *Trusters*. They defend civil rights above all, and they somehow entertain erosion of trust in the vaccines. Their stronger values for Moral Foundations are for Subversion and Harm.

The hesitancy profiles derived for the COVID-19 vaccine are detailed in Table 12, which also lists five profiles, along with the number of users in each hesitancy profile, defined by prototypical vector user representations. The interpretation of the five Twitter hesitancy profiles for the COVID-19 vaccines illustrated in Table 12 follows. The *Promoters* mostly trust in the role of the role of the COVID-19 vaccines for public health and believe that mandatory or expanded vaccination programs are needed. These users also are rejecting the “Lack of Trust in Safety of COVID-19 Vaccines”. These users focus on the Moral Foundations of Care, Authority, Loyalty, and Fairness and actively reject Subversion. The much larger group who make up the *Ambivalent* adopt some misinformation and are ambivalent in their trust in the vaccines – accepting and rejecting trust in the COVID-19 vaccines. Their beliefs in the civil rights when it comes to vaccination is also ambivalent, as they generally trust the safety and role of vaccines, but do not necessarily think the COVID-19 vaccines should be mandatory. These users focus on Authority, Harm, and Fairness.

**Table 12:** Twitter Hesitancy Profiles for the COVID-19 Vaccine.

|  |  |
| --- | --- |
| **Profile 1: PROMOTERS** (48,447 users; 9%)<br><u>Misinformation Themes:</u><br>-0.07: Unnecessary COVID-19 Vaccine<br>-0.07: Testing of COVID-19 Vaccine<br>-0.07: Adverse Events of COVID-19 Vaccine<br>+0.06: COVID-19 Vaccine Ingredients<br>-0.04: Unsafe COVID-19 Vaccine \| -0.04: Effect on Immune System<br>-0.03: Not effective COVID-19 Vaccine<br><u>Themes for Eroding Trust in Vaccine:</u><br>+0.35: De-Motivation in Taking the COVID-19 Vaccines<br>-0.10: Lack of Trust in Safety of COVID-19 Vaccines<br><u>Themes for Building Trust in Vaccine:</u><br>+1.03: Trust in Role of COVID-19 Vaccines for Public Health<br>+0.83: Motivation for Taking COVID-19 Vaccines<br>+0.22: Trust in the Safety of COVID-19 Vaccines<br>+0.05: Trust in Effects of COVID-19 Vaccines<br><u>Vaccine Literacy:</u> +2.32: Having literacy \| 0.04: Lacking literacy<br><u>Civil Rights:</u> +0.56: Vaccines more important than civil rights<br><u>Moral Foundations:</u><br>+1.63: Care \| +1.82: Authority \| +0.71: Loyalty \| +0.98: Fairness \|<br>-0.37: Subversion \| 0.64: Harm \| 0.03: Degradation \| 0.05: Purity | **Profile 2: AMBIVALENTS** (267,087 users; 48%)<br><u>Misinformation Themes:</u><br>+0.08: Information about COVID-19 Vaccines is Concealed<br>+0.07: COVID-19 Vaccine Ingredients<br>+0.03: Unsafe COVID-19 Vaccine<br><u>Themes for Eroding Trust in Vaccine:</u><br>+0.08: De-Motivation in Taking the COVID-19 Vaccines<br>+0.04: Lack of Trust in Safety of COVID-19 Vaccines<br>+0.03: Lack of Trust in Effects of COVID-19 Vaccines<br><u>Themes for Building Trust in Vaccines:</u><br>+0.08: Trust in Role of COVID-19 Vaccines for Public Health<br>+0.06: Trust in the Safety of COVID-19 Vaccines<br>+0.05: Motivation for Taking COVID-19 Vaccines<br><u>Vaccine Literacy:</u><br>+0.26: Having literacy<br><u>Civil Rights:</u><br>+0.27: Civil rights above all \|<br>+0.17: Vaccines more important than civil rights<br><u>Moral Foundations:</u><br>+0.15: Care \| +0.45: Authority \| +0.34: Fairness \|<br>+0.17: Loyalty \| +0.10: Betrayal \| +0.26: Subversion \|<br>+0.36: Harm \| +0.08: Degradation |
| **Profile 3: DEBUNKERS** (199,014 users; 35%)<br><u>Misinformation Themes:</u><br>-0.11: Unnecessary COVID-19 Vaccine<br>-0.06: Unsafe COVID-19 Vaccine \| -0.05: Testing of COVID-19 Vaccine<br>-0.05: Information about COVID-19 Vaccines is Concealed<br>-0.03: Effect on Immune System<br>-0.03: Adverse Events of COVID-19 Vaccine<br><u>Themes for Eroding Trust in Vaccine:</u><br>-0.27: Lack of Trust in Role of COVID-19 Vaccines for Public Health<br>-0.12: Lack of Trust in Safety of COVID-19 Vaccines<br>-0.07: Lack of Trust in the Role of Pharmaceutical Companies in Fighting COVID-19 Infections<br>-0.03: Lack of Trust in Effects of COVID-19 Vaccines<br><u>Vaccine Literacy:</u> -0.04: Having literacy<br><u>Civil Rights:</u> -0.26: Civil rights above all<br><u>Moral Foundations:</u><br>-0.35: Degradation \| -0.80: Subversion \| -0.09: Authority \| -0.11: Purity \|<br>-0.52: Harm \| -0.09: Care \| -0.07: Cheating \| -0.09: Betrayal | **Profile 4: MISINFORMERS** (5,617 users; 1%)<br><u>Misinformation Themes:</u><br>+0.83: Information about COVID-19 Vaccines is Concealed<br>+0.57: Testing of COVID-19 Vaccine<br>+0.41: COVID-19 Vaccine Ingredients<br>+0.40: Effect on Immune System<br>+0.28: Adverse Events of COVID-19 Vaccine<br>+0.27: Unsafe COVID-19 Vaccine<br>+0.25: Not effective COVID-19 Vaccine<br>+0.23: Unnecessary COVID-19 Vaccine<br>+0.19: Alternatives to COVID-19 Vaccine<br><u>Themes for Eroding Trust in Vaccine:</u><br>+1.54: De-Motivation in Taking the COVID-19 Vaccines<br>+0.85: Lack of Trust in Safety of COVID-19 Vaccines<br>+0.41: Lack of Trust in the Role of Pharmaceutical Companies in Fighting COVID-19 Infections<br>+0.36: Lack of Trust in Effects of COVID-19 Vaccines<br>+0.31: Lack of Trust in Role of COVID-19 Vaccines for Public Health<br>+0.20: Lack of Trust in Testing of COVID-19 Vaccines<br><u>Themes for Building Trust in Vaccine:</u><br>-0.13: Trust in the Safety of COVID-19 Vaccines<br>-0.10: Trust in Role of COVID-19 Vaccines for Public Health<br>-0.07: Trust in Effects of COVID-19 Vaccines<br>-0.07: Motivation for Taking COVID-19 Vaccines<br>+0.03: Trust in the Role of Pharmaceutical Companies in Fighting COVID-19 Infections<br><u>Vaccine Literacy:</u><br>+0.57: Having literacy \| +0.55: Lacking literacy<br><u>Civil Rights:</u><br>+2.46: Civil rights above all<br>+0.53: Vaccines more important than civil rights<br><u>Moral Foundations:</u><br>+1.93: Betrayal \| +4.82: Harm \| +1.68: Authority \| -0.06: Care<br>+0.67: Cheating \| +4.90: Subversion \| +0.98: Degradation \|<br>+2.02 Fairness \| +0.87 Loyalty \| +0.52 Purity |
| **Profile 5: SKEPTICS** (41,044 users; 7%)<br><u>Misinformation Themes:</u><br>+0.20: Information about COVID-19 Vaccines is Concealed<br>+0.06: COVID-19 Vaccine Ingredients \| +0.04: Adverse Events<br>+0.05: Testing of COVID-19 Vaccine \| +0.04: Effect on Immune System<br>+0.04: Not effective vaccine \| +0.04: Unnecessary COVID-19 Vaccine<br><u>Themes for Eroding Trust in Vaccine:</u><br>+1.31: De-Motivation in Taking the COVID-19 Vaccines<br>+0.07: Lack of Trust in Safety of COVID-19 Vaccines<br>+0.04: Lack of Trust in the Role of Pharmaceutical Companies in Fighting COVID-19 Infections<br>+0.03: Lack of Trust in Testing/Effects of COVID-19 Vaccines<br><u>Vaccine Literacy:</u> +0.23: Lacking literacy \| -0.09: Having literacy<br><u>Civil Rights:</u> +0.67: Civil rights above all<br><u>Moral Foundations:</u><br>+1.68: Subversion \| +1.00: Harm \| +1.37: Betrayal \| +0.11: Cheating \|<br>+0.14: Degradation \| +0.06: Purity \| +0.20: Fairness \| +0.31 Authority |  |

The COVID-19 *Misinformers* actively propagate varied misinformation about the COVID-19 vaccines and extremely oppose vaccination mandates. They also sow distrust in the COVID-19 vaccines. Their predominant Moral Foundations are Harm and Subversion. The *Skeptics* are Twitter users that while considering some misinformation, they seem to be mostly demotivated in vaccinating against COVID-19. Notably, these users generally lack vaccine literacy and are strong believers that their civil rights should be respected. Their predominant Moral Foundations are Subversion, Betrayal and Harm. Finally, *Debunkers* who comprise 35% reject misinformation, erosion of trust framings, civil rights above all, and reject a moral framework in their stance toward vaccine confidence framings. All the profiles discovered for HPV and COVID-19 vaccines and their interpretations provide answers to RQ5.

## DISCUSSION

The decision to discover framings of confidence in vaccines instead of working directly with tweets that discuss the HPV or the COVID-19 vaccines is rooted in our belief that while some tweets may be relevant to the questions from the Vaccine Confidence Repository (VCR), introduced in (Rossen et al., 2019), those tweets do not provide an explanation for *why* the tweet’s author agrees or disagrees with any of the VCR questions. Instead, framings, which were inferred from groups of tweets sharing the same attitude towards a specific question, provide the rationale behind agreeing or disagreeing with the VCR questions. Qualitatively, vaccine confidence for both HPV and COVID19 was expressed in framings covering a range of themes from general vaccine safety issues to individual level concerns (unsafe, adverse effects, ingredients, overwhelming the immune system), to vaccine development, testing and transparency concerns, to questioning vaccine efficacy, whether vaccinating is necessary, and alternatives. We therefore uncover not only vaccine confidence themes on social media, which may have been recognized in prior literature (Dunn et al., 2017; Islam et al., 2020; Shapiro et al., 2017; Wawrzuta et al., 2021), but importantly, uncover users’ stance toward those vaccine confidence themes and moreover, across millions of users at scale.

Quantitatively, we inferred a larger number of confidence framings for the COVID-19 vaccine (113 framings) than for the HPV vaccine (64 framings), perhaps because we operated on a larger number of tweets in the 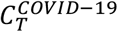 collection (5,865,046 unique tweets), which is an order of magnitude larger than the number of tweets in the collection 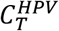 collection (422,078 unique tweets). But we also believe that the quantitative differences may be explained by the question/answering framework we designed to find the vaccine confidence framings, illustrated in Figure 1. We noticed that the experts judged a larger number of tweets relevant to the COVID-19 vaccine questions than they did for the HPV vaccine questions. This provides a second, and perhaps better explanation for why we obtained a different number of framings between the two vaccines, highlighting the finding that in Twitter discourse, people have a greater number of vaccine confidence issues for the COVID-19 vaccine than for the HPV vaccine.

The confidence framings that we collected in *framings*^*COVID*-19^ and *framings*^*HPV*^ allowed us to discover not only the fact that the distribution of framing categories varies between the vaccines, but also the fact in Twitter discourse about vaccines, vaccine confidence is impacted not only by misinformation, but also by the erosion of trust in vaccines. Some of the questions from VCR invite confidence framings that rely on misinformation. For example, Q11: “*Homeopathic medicines are an effective alternative to conventional vaccines*” produces as answers only misinformation framings, as shown in Figure 3. However, other questions, such as Q9: *“The more people who get vaccinated the greater the protection against disease”* are answered by framings that either build or erode trust in vaccines in the Twitter discourse about the HPV vaccine, clearly showcasing vaccine literacy problems. In the discourse about COVID-19 vaccination, the same question is answered by framings that only build trust in the vaccine, making use of vaccine literacy. Not surprisingly, framings answering the question Q13: “*It is important that people are able to make their own decisions about vaccination*” are dominated by civil rights issues for both vaccines. Another interesting observation derived from the analysis of Figure 3 is that framings relying on misinformation were inferred as answers to 11 questions when considering the HPV vaccine, while for the COVID-19 vaccine, misinformation was present in the framings answering 14 questions. Hence misinformation framings play an important role in answering more VCR questions for the COVID-19 vaccine.

While much interest has been shown in identifying misinformation on social media platforms, relatively few studies have considered addressing the problem of identifying the specific misinformation that is propagated on Twitter or other social media platforms (Luo et al., 2019; Margolis et al., 2019; Massey et al., 2020; Reiter et al., 2018). Typically, *known* misinformation can be identified, through methods such as (Weinzierl & Harabagiu, 2021) by relying on Wikipedia web pages or similar sources that collect debunked specific misinformation. However, our method of finding vaccine confidence framings as a Q/A problem produced an interesting byproduct, namely framings that contained specific misinformation, which we further analyzed to organize in misinformation taxonomies specific to the HPV or the COVID-19 vaccines. It is also important to note that the two misinformation taxonomies comprise typologies that discovered a greater number of misinformation themes than the typology of misinformation reported in Jamison et al (2020), which was adapted from Kata’s ontology (2010, 2012). Secondly, we also discerned concerns that sub-categorize each theme. In contrast, the taxonomy reported in (Jamison et al., 2020) is centered on pro or antivaccination sentiment, across all vaccines, whereas the misinformation taxonomies we derived were concerned only with confidence in specific vaccines. Perhaps the most interesting aspect of the comparison of the two misinformation taxonomies stems from the observation that although the framings were vastly different, and 90% of the themes were identical, whereas only 21% of the concerns specializing the themes in the taxonomies were shared. This indicates that misinformation that concerns vaccine confidence addresses 9 themes, e.g. vaccine testing, ingredients, vaccine alternative, etc, while the concerns that are used to develop the misinformation themes are vaccine-specific, and so are the framings. This accounts for the generative power of *Misinformers* and the tailoring of the misinformation to vaccines. But vaccine confidence is not impacted only by misinformation, as we have seen in Figure 3.

From the *framings*^*HPV*^, 21.8% increased trust in the safety of vaccines (n=21/96) while 20.8% of frames eroded trust in the safety of the HPV vaccine (N=20/96). From the *framings*^*COVID*-19^, 24.1% increased trust in the safety of the vaccines (n=27/112) while 22.3% eroded trust in the safety of vaccines (n=25/112). This motivated our decision to derive two different trust taxonomies for each vaccine: one for eroding trust, the other for building trust in the vaccines. To our knowledge, this is the first time when trust in vaccines has considered either the erosion of trust or the increase in trust. Moreover, the empirically derived taxonomies of trust in the COVID-19 and HPV vaccines reveal a number of findings. First, Twitter discourse on vaccination expressed vaccine attitudes in relation to trust to a substantial degree (20-24% of vaccine stance framings) and in approximate equal proportions in relation to building or eroding trust across vaccines. Second, trust in vaccines was expressed across both HPV and COVID19 vaccines at the individual level (e.g., confidence in vaccine over natural immunity), the family level (e.g., vaccination protects families), to the system level (e.g., vaccine prevents cancer regardless of pharma profit, government provides and makes transparent vaccine information). Thus, a social-ecological framework contextualized expressions of trust at multiple levels on twitter (Latkin et al., 2021).

Common arguments across vaccines that were grounded in *building* trust included four areas: general statements asserting vaccine safety, government transparency about adverse effect reporting, vaccine immunity is better than natural immunity, and that ineffective vaccines are not profitable for pharmaceutical companies. For expressions of eroding trust, one concern was shared across vaccines namely, lack of trust in pharmaceutical companies – that they produce ineffective vaccines for profit, and this is evidenced by them producing and recommending annual boosters. Equal numbers of arguments at the individual and system levels of trust were expressed in both building and eroding trust across vaccines.

Qualitatively, trust was discussed in terms of general statements asserting that vaccination is safe and effective, to safety information being publicly available, to motivations to vaccinate that include avoiding cancer or COVID19 which is worse than vaccine side effects, vaccination triggering a more reliable immune response, to vaccinating children to be able to return to in-person school (in the case of COVID19), prioritizing vaccination as key to controlling infectious disease, protecting others, to mRNA leaves your body soon after you vaccinate. Trust in doctors and scientists explaining the benefits of vaccinating was present among HPV vaccine tweets only, but not COVID-19 surprisingly.

Erosion of trust was expressed by instilling doubt in vaccination by stating it is unknown whether the COVID vaccine protects against future variants, to vaccines could worsen existing health conditions, to individuals with allergies should not vaccinate, to explicitly claiming the vaccines are experimental, that long term side effects are not known and more time is needed, to pharmaceutical companies profiting at the expense of health of individuals, the vaccine development process was rushed, children should not be vaccinated because there is no legal accountability for adverse events, to more proof is needed and questioning vaccine ingredients (e.g., aluminum). Again, only for HPV vaccination were claims made that doctors and scientists do not tell the truth about vaccines with explicit doubt was cast on published research. These themes were also recognized in recent research on COVID-19 vaccination and trust by Latkin et al (2021). Trust in the vaccine literature has commonly been measured with single items (Larson et al., 2018), has often been operationalized by measuring credibility source (Larson et al., 2018; Sutton et al., 2020) but has rarely been operationalized or measured as perceived motivations or erosion of trust or as moral values that align. Only recently has one trust scale been developed that measures parental confidence in source credibility and trust in various sources but also measures the norms that vaccination is important for children, a protective measure that all teenagers should get vaccinated (Frew et al., 2019). These trust expressions on twitter however it is important to remember that they are unsolicited trust expressions as opposed to survey results. Twitter framings that erode trust focus on instilling fear or casting doubt, de-motivating vaccination at the individual level, but also focusing attention on the failure and incompetence of institutional systems and experts. This has been highlighted in Recreancy and social capital theory, which explains loss of trust and credibility in contentious public health disasters by emphasizing institutional failures of responsibility (Freudenburg, 1993).

It is important to note our usage of the implied Moral Foundations (MFs) is also new in its application to understanding vaccine confidence and hesitancy expressions on social media. More importantly, that we have associated each implied MFs with a *stance*. The stance of each author of a tweet towards the framing (s)he refers is transferred to the MFs implied by the same framing. Based on this observation, clear moral attitudes emerge within the hesitancy profiles across both the HPV and COVID-19 vaccines. *Promoters* of both the HPV and COVID-19 vaccines tend to strongly accept framings, which espouse Care, Authority, Loyalty, and Fairness, with rejection of Subversion. Alternatively, *Misinformers* of both the HPV and COVID-19 vaccines tend to adopt framings in stark moral contrast than *Promoters*, whose moral foundations of Betrayal, Harm, and Subversion oppose those moral stances of Loyalty, Care, and Authority respectively. *Misinformers* tend to have much stronger moral stances than *Promoters*, which indicates morality plays a key role in the motivation of those spreading misinformation at scale. A similar pattern is found when comparing the *Skeptic* profiles, where moral foundations of Subversion, Betrayal, and Harm are adopted towards both the HPV and COVID-19 vaccines. In contrast, we find slightly differing moral profiles across vaccines when comparing the *Trusters* of the HPV Vaccine to the *Ambivalent* of the COVID-19 Vaccine. The two groups share in their adoption of Authority and Fairness moral foundations, but the HPV Vaccine *Trusters* adopt Care, while the COVID-19 Vaccine *Ambivalent* adopt Harm. The *Debunkers* share in rejection of Subversion but differ across other moral foundations. HPV Vaccine *Debunkers* equally tend to reject Degradation, Subversion, and Authority, while COVID-19 Vaccine *Debunkers* focus much more on rejecting Subversion, with a secondary focus on Harm and Fairness.

While the vector user representations that we learned enabled us to discover the profiles listed in Tables 11 and 12, the interpretations of the hesitancy profiles require additional discussions. Five vaccine hesitancy profiles were identified for each vaccine as the optimal number of profiles that distinguished users who expressed a stance toward vaccine framings for HPV and COVID19. These vaccine profiles reflect subgroups who reject or accept various vaccine confidence framings, which were further qualified across six categories: (1) misinformation exposure; (2) building trust, eroding trust; (4) Moral Foundations with ten possible values (care/harm; authority/subversion; fairness/cheating; loyalty/betrayal; purity/degradation), (5) civil rights (preference for mandating vaccines or civil rights above all irrespective of public health circumstances); and (6) vaccine literacy. Of note is that HPV vaccine profiles reflect stance toward framings derived across thirteen years from 2008 through 2021 while COVID-19 profiles reflect stance toward framings derived from tweets users spanning 7 months from January through July 2021 in the context of the pandemic.

## VACCINE HESITANCY PROFILES AS PERSON CENTERED AUDIENCE SEGMENTATION FOR TARGETED CAMPAIGNS

HPV and COVID19 vaccine hesitancy profiles highlight a constellation of accept and reject stances across various framings, which will inform future messaging campaigns. The potential range of messaging targets spans inoculating against specific misinformation to tapping into moral frameworks to importantly, ways to bolster trust or debunk messaging that erodes trust. Vaccine stance identified from social media importantly, should be distinguished in its value for reflecting unsolicited attitudes toward vaccines in contrast to survey research (Hornik et al., 2020). Although five profiles were discerned for each vaccine, there are substantial differences both quantitatively in the relative size of profiles, and qualitatively, how profiles distinguish users.

Interest to public health interventionists involves strategically targeting profile members whose stance suggest their vaccine attitudes are amenable to change, or alternatively, whose vaccine attitudes may already be positive but need strengthening (Roskos-Ewoldsen et al., 2002). With this goal in mind, *Promoters* (21%) and *Debunkers* (32%) who make up more than half of the HPV vaccine users, express to a large degree support for vaccination in their high motivation to vaccinate, their trust, being vaccine literate, supportive of mandating vaccination, and appeal to moral frames of care, authority and loyalty in the case of *Promoters*. These users may respond to authoritative appeals, mandating vaccination, and trust appeals that emphasize the importance of public health. For COVID-19 vaccine profiles on the other hand, whose stance may also already be positive but in need of strengthening, *Promoters* make up a much smaller subgroup (9%) while *Debunkers* make up 35%. Bolstering positive vaccine attitudes may be achieved with trust messaging that emphasizes the importance of vaccinating for public health i.e., the collective, and motivating vaccination by emphasizing moral values of care (preventing harm), authority, loyalty, and fairness for Promoters. By contrast, for *Debunkers* – who make up a substantial subgroup (almost 200,000 users), morality messaging should be avoided with this subgroup who reject moral framings. An emphasis on moral messages with this subgroup may boomerang (Fishbein et al., 2002).

Of greater interest are profiles whose frame-stance scores suggest these profile users are ambivalent, on-the-fence, or skeptical whose members are more likely to be unvaccinated and hold vaccine attitudes amenable to change. Among HPV vaccine profiles, *Trusters* comprised the second largest subgroup (28%) after *Debunkers* and make up a substantial group in size relatively speaking (nearly 20,000). The pool of HPV vaccine users overall was smaller than that of COVID19 vaccine users – representing a vastly larger population of Twitter users. *Trusters* are accepting of a range of vaccine themes that build trust (e.g., motivated to vaccinate, trust in the role of public health, doctors, science, and the effects of the vaccine). These findings on frame-stance score suggest that *Trusters* are likely to respond favorably to messages that build trust. These users are literate and their motivation to vaccinate can be tapped possibly through moral value appeals of care, authority and fairness. Avoiding messaging that emphasizes vaccine mandates is warranted for this subgroup given *Trusters*’ weak yet existing stance on civil rights above all irrespective of public health circumstances. Findings from our study therefore inform not only messaging that users may respond to but also messaging that should be avoided in order to prevent potential iatrogenic message effects (Fishbein et al., 2002; Moos, 2005).

In comparison to *Trusters* among HPV vaccine profiles, the *Ambivalent*, who make up the largest subgroup (48%) of COVID-19 vaccine hesitancy profiles with 267,087 users, also reveal a weak yet existing frame-stance score across trust themes in COVID-19 vaccination. These users truly are on the fence, both accepting and rejecting trust framings and whose motivation to vaccinate needs to be strengthened. This subgroup may benefit from significantly bolstering trust and motivation coupled with inoculating against misinformation and utilizing moral appeals of authority and preventing harm. Both *Trusters* (HPV) and the *Ambivalent* (COVID19) are ripe for receiving inoculation messages against misinformation across vaccine safety, effectiveness, the testing process, transparency, ingredients, and adverse reactions. Similar misinformation domains have been recognized in the literature across HPV and COVID-19 yet never in this social media vaccine frame-stance context at this scale (Calo et al., 2021; Head et al., 2018; Jamison et al., 2020; Kim et al., 2020; Loomba et al., 2021; Massey et al., 2020; Sundstrom et al., 2021; Van der Linden et al., 2016; Zimet et al., 2013).

A smaller subgroup of vaccine profiles, the *Skeptics* (13% for HPV; 7% for COVID19), exhibit frame-stance scores that are accepting of most misinformation and erosion of trust framings. These users are illiterate, strongly de-motivated to vaccinate, and with whom moral values of subversion, harm and betrayal resonate as well as civil right above all irrespective of public health circumstances. Reaching these users presents more challenges. These subgroups’ vaccine stances are not as extreme as *Misinformers* who widely distrust vaccination and actively propagate misinformation. The *Skeptics* exhibit weak frame-stance scores on many fronts suggesting these could be targeted to shift vaccine attitudes.

## STENGTHS AND LIMITATIONS

Strengths of the methodology for discovering vaccine confidence framings presented in this paper include (1) the Q/A framework that was used as a starting point to identify framings; (2) the inference of framings that answer questions from the Vaccine Confidence Repository (VCR); (3) the discovery at scale of tweets that refer to the confidence framings and (4) the identification of the stance of the tweets towards vaccine confidence framings. The Q/A framework pinpoints tweets that are relevant to the VCR questions, from which framings were inferred by a method inspired by multi-document summarization (Nenkova & Passonneau, 2004). These sophisticated natural language processing methods have not been used before in processing tweets discussing vaccine hesitancy. They have the advantage of operating at the pragmatic level of language processing, in contrast with the topic processing methods, which operate at the lexical level. Furthermore, the results of topic modeling methods are notorious to be difficult to interpret, whereas the framings are not only insightful, but also more straightforward to interpret. The framings are insightful because they enabled us to identify misinformation, trust in vaccines, civil rights and morality issues that were discussed. The framings also revealed the vaccine literacy of Twitter users. Moreover, by discovering framings at scale, this method considers the viewpoint of 138,779 Twitter users regarding their confidence in the HPV vaccine (i.e., users who specifically expressed their stance toward HPV and COVID-19 vaccine confidence) and of 665,798 Twitter users regarding their confidence in the COVID-19 vaccines. By identifying the stance of tweets towards each framing we have used a more advanced form of affect processing of the language in Tweets than the one afforded by sentiment analysis. This is because sentiment analysis operates at the lexical level, considering the positive, negative, or neutral orientation of words to infer the sentiment of a tweet. In contrast, stance identification considers the interaction of lexical, syntactic and semantic features of a tweet’s language with emotions to derive the attitude of a tweet towards a specific framing and results in the identification of subgroups of *users*, which is informative to a greater degree for public health campaign design. For example, a tweet may have positive sentiment, but its stance may be rejecting a given framing.

The discovery of the hesitancy profiles, which is unique to the method presented in this paper, is another notable strength, made possible by (a) the recognition of *framings*^*HPV*^ and *framings*^*COVID*-19^ at scale; (b) the identification of the stance of tweets referring to any of these framings and (c) the representation of the users authoring these tweets though vector user representations (VURs) encoding values informed by their stance in various confidence framings, comprising misinformation they accepted or rejected, their trust in vaccines and role of civil rights, their vaccine literacy as well as their implied Moral Foundations. Perhaps the most important strength of the way in which users have been represented stems from the usage of the themes of the misinformation and trust taxonomies to account for the values the users put on the framings they referred to.

The method described in the paper has also some important limitations. First, we do not know if *framings*^*HPV*^ and *framings*^*COVID*-19^ represent all the framings referred on Twitter about vaccine confidence in the two types of vaccines. To address this issue, we would need to consider additional questions that address vaccine confidence and find if new framings were inferred for the new questions. Second, the framings were inferred from tweets deemed relevant to the questions by inspecting the top-ranked 300 tweets. As in all information retrieval tasks, recall of all relevant tweets from 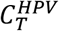 or 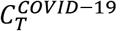 is difficult to achieve without reading all tweets in the collections, which is an enormous task. Because the confidence framings are central to the method described in this paper, we believe that the Misinformation and Trust Taxonomies also have the same limitation of completeness, which also impacts on the completeness of the hesitancy profiles. A limitation of the hesitancy profiles that were discovered is also determined by our exclusive focus on vaccine confidence, while hesitancy should also account for vaccine convenience and complacency.

While the interpretation of the hesitancy profiles is insightful, future work will need to test and validate these profiles for both user vaccination status and for profile member responsiveness to strategic messaging. Six vaccine relevant value frameworks characterize these vaccine profiles for HPV and COVID-19 vaccination – two voluntary and underutilized vaccines shown to be safe and effective. Derived from millions of tweets and unsolicited vaccine attitudes expressed on Twitter - these factors contribute as first steps to identify and characterize the complexity in vaccine hesitancy profiles at such scale. The implication with these profiles indicates promise to reach vastly larger number of unvaccinated with more precise and strategically targeted messaging. The misinformation and trust taxonomies that informed the vaccine profiles shed light on nuanced differences and similarities among subgroups and between vaccines in regard to stance on moral foundations, trust, and misinformation dimensions that contribute to vaccine attitudes and importantly informs which vaccine attitudes may be accessible and amenable to change for each subgroup. Prior research has demonstrated the importance of considering each of the five value frameworks applied in this study to characterize vaccine stance profiles. Loomba et al (2021) as one example, were the first to quantify the impact of misinformation exposure on vaccine hesitancy.

## CONCLUSION

In this paper we have presented a method capable of inferring 64 ways in which confidence in the HPV vaccine is framed in a collection of 422,078 unique tweets and 113 ways in which confidence in the COVID-19 vaccines is framed in a collection of 5,865,046 unique tweets. Moreover, these confidence framings inform a taxonomy of misinformation in HPV vaccines as well as a taxonomy of misinformation in the COVID-19 vaccines, providing insights into the themes of misinformation in these vaccines that are propagated on Twitter, as well as the concerns that are addressed with misinformation. Furthermore, the confidence framings informed taxonomies indicating how trust in the vaccines is eroded as well as how trust in the vaccine is increased in the Twitter discourse. Using these taxonomies along with analyzing the vaccine literacy, the implied moral foundations of each framing and the tension between vaccine mandates and civil rights allowed us to discover several profiles of hesitancy for each vaccine. The discovery of these profiles was made possible by (a) the automatic recognition of all tweets from the entire tweet collection that refer to some of the 64 framings of confidence in the HPV vaccine and the similar automatic recognition of all tweets from the entire tweet collection that refer to some of the 113 framings of confidence in the COVID-19 vaccine; and (b) the automatic identification of the stance these tweets have towards the referred framings.

This novel methodology sheds light on what has been known but rarely modelled in this detailed, in-depth manner, namely the heterogeneity that makes up vaccine attitude profiles. Furthermore, this novel modeling approach captures user stance toward vaccine framings that uncovers the attitude orientation and informs messaging that can tap into which vaccine attitudes may be accessible (Roskos-Ewoldsen et al., 2002). Processing the language of vaccine confidence, coupled with stance discovery toward dimensions of vaccine confidence, misinformation, erosion and building of trust, literacy, civil rights, and five core moral value frameworks allowed for the discovery of nuanced rich vaccine hesitancy profiles that were discovered at very large scale of Twitter users. These results begin to disentangle the complex attitudes shaping vaccine attitudes. Furthermore, such a person or user-centered approach to characterizing vaccine hesitancy profiles recognizes the importance of uncovering subgroups with similar vaccine stance across multiple values. The patterns of vaccine framings across multiple value frameworks inform public health messaging approaches to effectively reach profiles with promise to shift or bolster vaccine attitudes.

## Data Availability

Twitter forbids the release of raw twitter data, but annotations and taxonomies will be released upon acceptance.

## SUPPLIMENTARY MATERIAL

**Table 13:** HPV Vaccine Confidence Questions used to discover framings.

| Question Id | Question Text |
| --- | --- |
| Q1 | How confident are you in the safety of the HPV vaccine |
| Q2 | Vaccines have not been adequately tested for safety. |
| Q3 | Vaccines overwhelm a child's undeveloped immune system |
| Q4 | Vaccines can cause or worsen allergies |
| Q5 | Vaccines introduce unnatural toxins into the body |
| Q6 | Getting vaccinated helps protect those who are unable to be vaccinated against disease |
| Q7 | Infectious diseases are virtually eliminated so vaccination is not needed. |
| Q8 | Vaccines cause the diseases they are supposed to prevent |
| Q9 | The more people who get vaccinated the greater the protection against disease |
| Q10 | Improved living standards, not vaccination, have reduced infectious diseases. |
| Q11 | Homeopathic medicines are an effective alternative to conventional vaccines |
| Q12 | Building immunity by naturally fighting off a disease is better protection than getting a vaccine. |
| Q13 | It is important that people are able to make their own decisions about vaccination. |
| Q14 | It should be compulsory for all children to be vaccinated. |
| Q15 | It is okay for people to be exempt from vaccination for moral or personal reasons. |
| Q16 | People should be able to decide whether or not to vaccinate their children. |
| Q17 | Pharmaceutical companies create ineffective vaccines in order to make high profit. |
| Q18 | The government conceals information about the safety of vaccines. |
| Q19 | Pharmaceutical companies purposefully conceal information about the safety of vaccines. |

**Table 14:** COVID-19 Vaccine Confidence Questions used to discover framings.

| Question Id | Question Text |
| --- | --- |
| Q1 | How confident are you in the safety of the COVID-19 vaccine |
| Q2 | Vaccines have not been adequately tested for safety. |
| Q3 | Vaccines overwhelm a child's undeveloped immune system |
| Q4 | Vaccines can cause or worsen allergies |
| Q5 | Vaccines introduce unnatural toxins into the body |
| Q6 | Getting vaccinated helps protect those who are unable to be vaccinated against disease |
| Q7 | Given all the COVID-19 variants, the protective effects of the vaccine do not last. |
| Q8 | If vaccinating against COVID-19, you will test positive. |
| Q9 | The more people who get vaccinated the greater the protection against disease |
| Q10 | Strong immune system, not vaccination, have reduced COVID-19 infections. |
| Q11 | Homeopathic medicines are an effective alternative to conventional vaccines |
| Q12 | Building immunity by naturally fighting off a disease is better protection than getting a vaccine. |
| Q13 | It is important that people are able to make their own decisions about vaccination. |
| Q14 | It should be compulsory for all children over 12 y/o to be vaccinated. |
| Q15 | It is okay for people to be exempt from vaccination for moral or personal reasons. |
| Q16 | People should be able to decide whether or not to vaccinate against COVID-19. |
| Q17 | Pharmaceutical companies create ineffective vaccines in order to make high profit. |
| Q18 | The government conceals information about the safety of vaccines. |
| Q19 | Pharmaceutical companies purposefully conceal information about the safety of vaccines. |

